# Gut community structure as a risk factor for infection in *Klebsiella*-colonized patients

**DOI:** 10.1101/2023.04.18.23288742

**Authors:** Jay Vornhagen, Krishna Rao, Michael A. Bachman

**Author notes:** Co-corresponding authors: Jay Vornhagen, Michael A. Bachman.

## Abstract

The primary risk factor for infection with members of *the Klebsiella pneumoniae* species complex is prior gut colonization, and infection is often caused by the colonizing strain. Despite the importance of the gut as a reservoir for infectious *Klebsiella*, little is known about the association between the gut microbiome and infection. To explore this relationship, we undertook a case-control study comparing the gut community structure of *Klebsiella*-colonized intensive care and hematology/oncology patients. Cases were *Klebsiella*-colonized patients infected by their colonizing strain (N = 83). Controls were *Klebsiella*-colonized patients that remained asymptomatic (N = 149). First, we characterized the gut community structure of *Klebsiella*-colonized patients agnostic to case status. Next, we determined that gut community data is useful for classifying cases and controls using machine learning models and that the gut community structure differed between cases and controls. *Klebsiella* relative abundance, a known risk factor for infection, had the greatest feature importance but other gut microbes were also informative. Finally, we show that integration of gut community structure with bacterial genotype or clinical variable data enhanced the ability of machine learning models to discriminate cases and controls. This study demonstrates that including gut community data with patient- and *Klebsiella*-derived biomarkers improves our ability to predict infection in *Klebsiella*-colonized patients.

**Importance:** Colonization is generally the first step in pathogenesis for bacteria with pathogenic potential. This step provides a unique window for intervention since a given potential pathogen has yet to cause damage to its host. Moreover, intervention during the colonization stage may help alleviate the burden of therapy failure as antimicrobial resistance rises. Yet, to understand the therapeutic potential of interventions that target colonization, we must first understand the biology of colonization and if biomarkers at the colonization stage can be used to stratify infection risk. The bacterial genus *Klebsiella* includes many species with varying degrees of pathogenic potential. Members of the *K. pneumoniae* species complex have the highest pathogenic potential. Patients colonized in their gut by these bacteria are at higher risk of subsequent infection with their colonizing strain. However, we do not understand if other members of the gut microbiota can be used as a biomarker to predict infection risk. In this study, we show that the gut microbiota differs between colonized patients that develop an infection versus those that do not. Additionally, we show that integrating gut microbiota data with patient and bacterial factors improves the ability to predict infections. As we continue to explore colonization as an intervention point to prevent infections in individuals colonized by potential pathogens, we must develop effective means for predicting and stratifying infection risk.

## Introduction

The gut is a vast ecosystem populated by trillions of bacteria, viruses, and microbial eukaryotes. The majority of these microbes have beneficial or neutral impacts on host health; however, some are potential pathogens. Under specific circumstances, some gut microbes can escape to distant body sites, leading to infection. One such group of pathogens is the *Klebsiella pneumoniae* species complex (referred to as “*Klebsiella*”). This complex contains several potentially pathogenic species of *Klebsiella*, including *K. pneumoniae*, *K. variicola*, *K. quasipneumoniae*, *K. quasivariicola* sp. nov., and *K. africana* (reviewed in (1)). These bacteria are common causes of bacteremia, pneumonia, and urinary tract infection (UTI). The genome content of a given strain of *Klebsiella* determines its infectious potential, where the presence of virulence and fitness factors permits and enhances infectivity, and antimicrobial resistance genes complicate infection treatment (2). As of 2019, *Klebsiella* is the third leading global cause of death attributable to, or associated with, antimicrobial resistance (3). More research is necessary to understand the *Klebsiella* pathogenesis. Such research may lead to improved diagnosis and treatment, and therein reduce the burden of *Klebsiella* disease.

*Klebsiella-*colonized patients are at increased risk for subsequent infection (4–6). Though few patient-centered studies determine the specific origin of infectious *Klebsiella*, those that have demonstrate that *Klebsiella-*colonized patients are infected with their colonizing strains in about ∼80% of cases (4, 6, 7). Additionally, gut dominance by *Klebsiella* is a risk factor for infection in *Klebsiella-* colonized patients (8–10). The identification and interrogation of factors that permit, enhance, or restrict *Klebsiella* gut colonization are receiving increased attention due to the clear importance of the gut as a reservoir for infectious *Klebsiella*. Recent laboratory-based studies have identified novel gut fitness factors (11–14), microbes that enhance colonization resistance (15, 16), and gut community structures that are permissive or restrictive to colonization (11, 17). Despite increased interest, studies aiming to understand gut ecology in *Klebsiella-*colonized patients are comparatively sparse, limiting the translatability of laboratory-based findings to real-world settings.

Previously, we performed a cohort study of over 1,900 *Klebsiella*-colonized patients in the intensive care and hematology/oncology units (7). The goal of this study was to identify patient variables associated with infection and two corresponding nested case-control studies were performed to assess the role of gut dominance in *Klebsiella* infection (8) and to rigorously identify infection-associated *Klebsiella* factors (18). Here, we aimed to leverage this case-control cohort of patients to understand the gut ecology of *Klebsiella-*colonized patients and determine if microbiome-derived biomarkers can improve infection prediction in *Klebsiella*-colonized patients.

## Results

### Description of study population

238 patients were originally selected from a cohort of 1,978 *Klebsiella* colonized intensive care and hematology/oncology patients (7) for a nested case-control study to assess the role of gut colonization density as a risk factor for *Klebsiella* infection (8). Cases were defined as colonized patients who met clinical criteria for infection (see prior publications for detailed criteria and physician case review process) with a *Klebsiella* strain that was detectable in the gut prior to infection. Controls had rectal colonization but no subsequent, symptomatic clinical infection. Cases were matched 1:2 with asymptomatically colonized controls based on rectal swab collection date, age, and sex. For the present study, we selected 232 patients (Table 1) from the previous study based on inclusion in our previous comparative genomics study and available DNA extracted from the rectal swab most proximal to the infection (18). The most common infection type was bacteremia, followed by UTI and respiratory infection (Table 1). 16S rRNA sequencing was performed using the method described by Kozich *et al.* 2013 (19).

**Table 1.** Select patient demographics

| VARIABLE |  | CASE<br>(N = 83) | CONTROL<br>(N = 149) | P-VALUE* |
| --- | --- | --- | --- | --- |
| AGE | mean ± SD | 60 ± 13 | 59 ± 12 | 0.556 |
| SEX | male | 43 (51.8%) | 76 (51.0%) | 1.000 |
|  | female | 40 (48.2%) | 71 (47.7%) |  |
|  | missing | 0 (0%) | 2 (1.3%) |  |
| RACE | white | 70 (84.3%) | 119 (79.9%) | 0.592 |
|  | nonwhite | 13 (15.7%) | 28 (18.8%) |  |
|  | missing | 0 (0%) | 2 (1.3%) |  |
| INFECTION SITE | blood | 41 (49.4%) |  |  |
|  | respiratory | 19 (22.9%) |  |  |
|  | urine | 23 (27.7%) |  |  |

\*age: student's *t* test; sex/race: fisher's exact test

### Description of the gut community of *Klebsiella-*colonized patients

First, we aimed to explore the gut community structure of *Klebsiella-*colonized patients agnostic of case status. *Klebsiella*, *Enterococcus*, *Escherichia/Shigella*, *Finegoldia*, and *Peptoniphilus* were the dominant gut microbiota in this study population (Figure 1A). Probabilistic modelling using Dirichlet multinomial mixtures (20) was used to determine if metacommunities exist in our study population. The optimal number of community clusters was two (Laplace approximation = 194340.97, Table S1), though one and three-community clusters yielded similar fits (one-community Laplace approximation = 194864.80, three-community Laplace approximation = 200401.69, Table S1). Case status was not associated with metacommunity structure in either the two (partition 1 v partition 2 odds ratio [95% CI] = 1.35 [0.789-2.32]) or three-community models (partition 1 v partition 2 odds ratio [95% CI] = 1.11 [0.505-2.46], partition 1 v partition 3 odds ratio [95% CI] = 0.921 [0.414-2.05], partition 2 v partition 3 odds ratio [95% CI] = 0.826 [0.46-1.48]). Principle coordinates analysis revealed that *Klebsiella* and *Enterococcus* were strong components determining metacommunity structure in both two (Partition 1, Figure 1B) and three-partition communities (Partition 3, Figure 1C), whereas other dominant gut microbiota influence different metacommunities (Figures 1B, C). Alpha-diversity analysis of these metacommunities revealed that *Klebsiella* influenced partitions (Partition 1 and Partition 3 in two and three partition communities, respectively) were significantly less rich (Chao), even (Shannon) and diverse (Inverse Simpson) than other metacommunities (Figures 1D-I). Interestingly, Partition 1 of the three-partition community clustering, which is heavily influenced by *Escherichia/Shigella* (Figure 1C), was significantly less rich, even, and diverse than Partition 2 (Figure 1G-I), which is influenced by *Finegoldia* and *Peptoniphilus* (Figure 1C). Given that *Finegoldia* and *Peptoniphilus* are strict anaerobes and *Klebsiella*, *Enterococcus*, and *Escherichia/Shigella* are facultative anaerobes, it may be the case that alpha diversity is driven by the presence or absence of anaerobic bacteria in the gut in this patient population. Collectively, these data indicate that *Klebsiella* is the dominant gut microbe in this population of *Klebsiella-*colonized patients, and is associated with reduced richness, evenness, and diversity.

**Figure 1.**
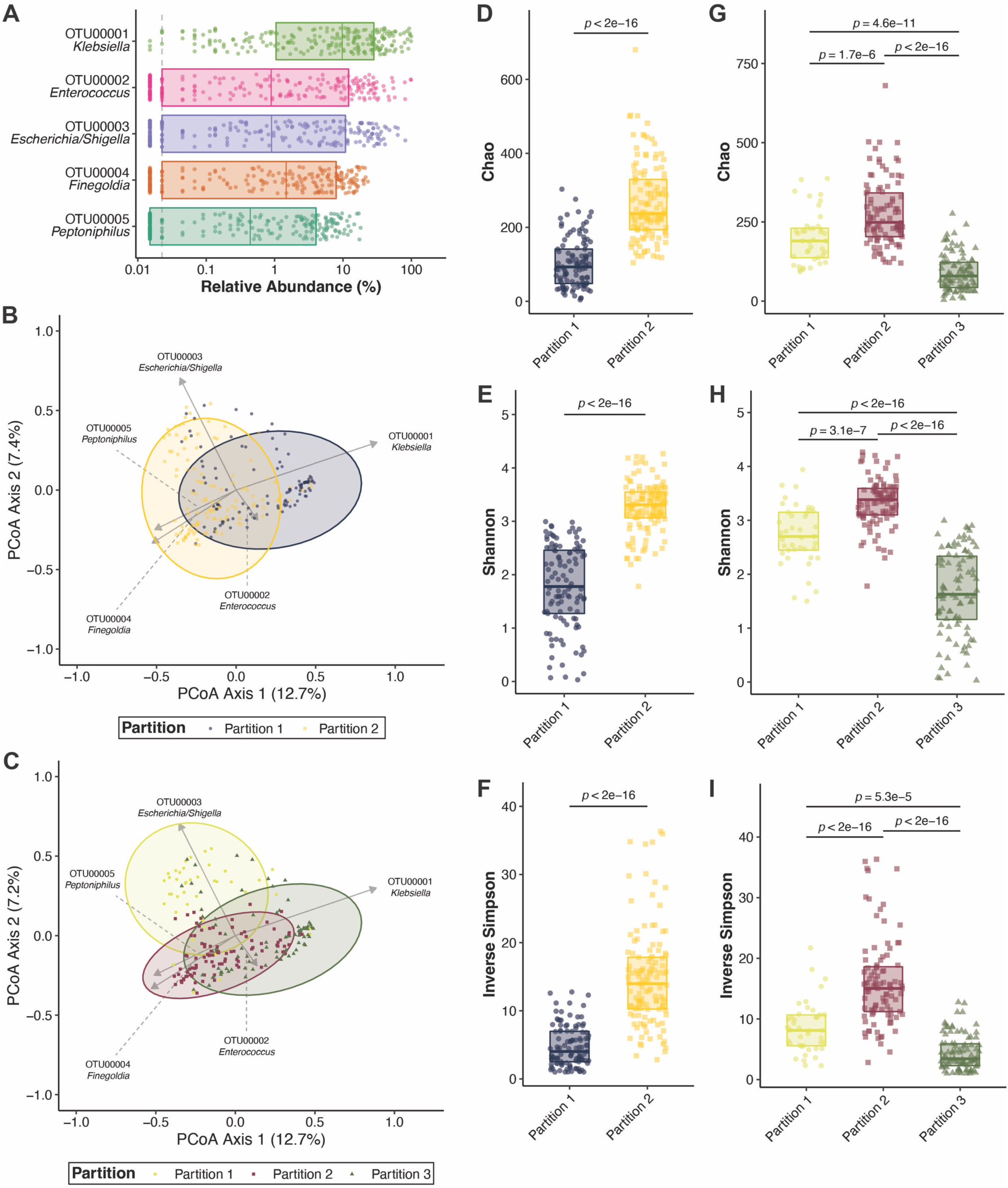
*Klebsiella* is the dominant gut microbe in *Klebsiella-*colonized patients. (A) Top five operational taxonomic units (OTUs) in *Klebsiella-*colonized patients (N = 232). Principal coordinates analysis with overlayed biplots of OTUs of two (B) and three-partition (C) community clustering using Dirichlet multinomial mixtures. Analysis of the Chao, Shannon, and Inverse Simpson alpha-diversity indices in two (D-F) and three-partition community clustering (G-I, boxplot indicates median with interquartile range, *p* indicates student’s *t* test *p-*value after Benjamini & Hochberg correction for multiple comparisons). For all panels, each datapoint indicates one patient.

### Determination of optimal taxonomic level to classify cases and controls

Out next goal was to determine the ability of microbiota composition to discriminate cases from controls. To this end, we used supervised machine learning models to classify case status, using different taxonomic levels as input data. Due to their high interpretability compared to other methods, we chose to use regularized logistic regression. To ensure optimal model performance, training was iterated across several combinations of hyperparameters (as in (21)), wherein the hyperparameter combination that yielded peak training performance was used for the final model (Figure S1). This process was repeated for phylum, class, order, family, genus, OTU, and amplicon sequence variant (ASV) level-data. ASVs provided the most robust discrimination of cases and controls (median area under the receiver-operator characteristic curve [AUC] = 0.68), followed by OTU (median AUC = 0.64) and phylum (median AUC = 0.63, Figure 2A). Additionally, models using ASVs as their input variables were most likely to yield an AUC > 0.5, indicating that classification of cases and controls was better than random chance. We found similar outcomes using the random forest method (Figure S2A), indicating that our results are robust across models that differ in method and interpretability. As we observed optimal model performance with ASVs, we decided to use the taxonomic level for the further study analyses.

**Figure 2.**
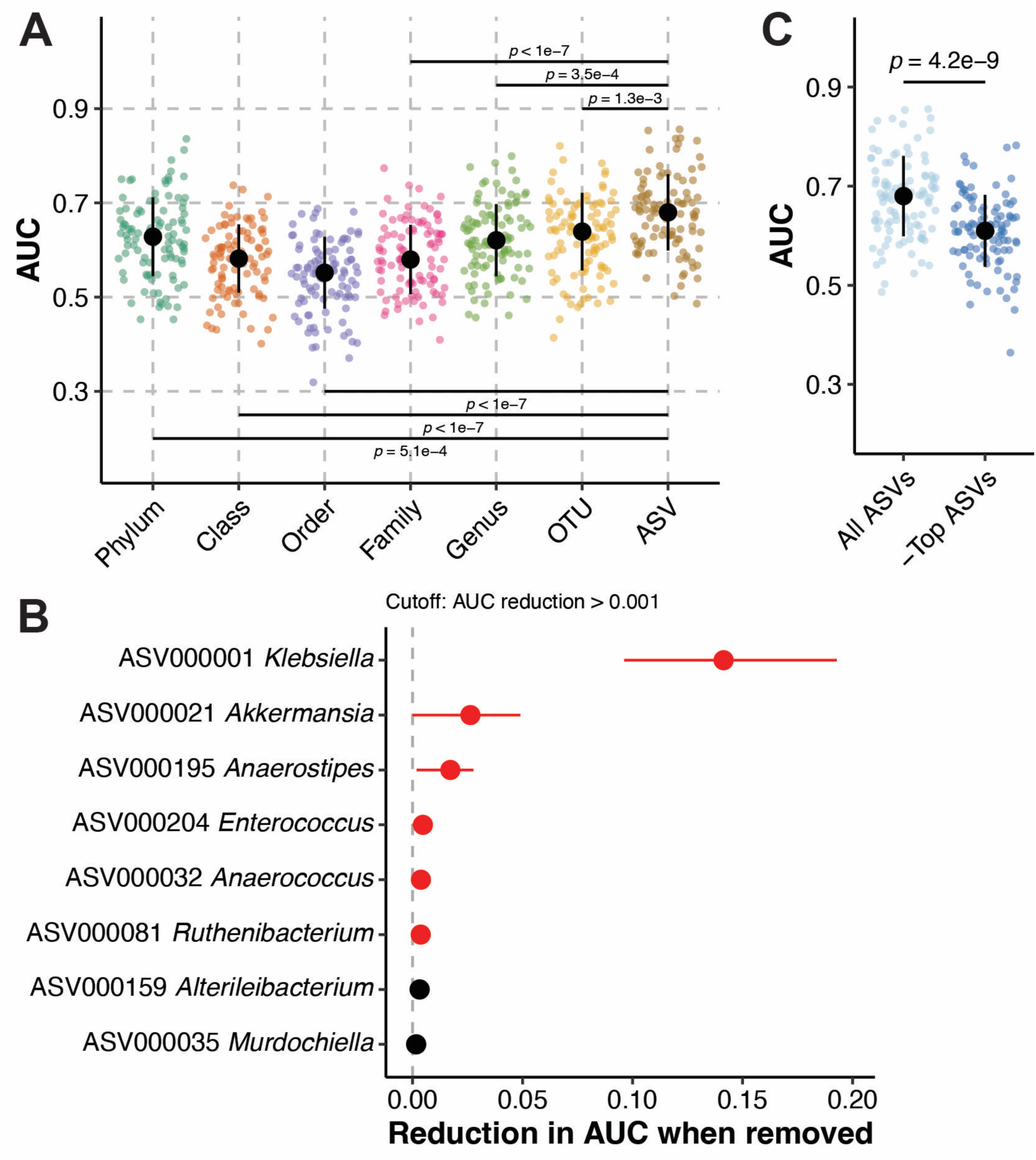
Amplicon sequence variants best discriminate cases and controls. (A) Regularized logistic regression model performance, as measured by area under the receiver-operator characteristic curve (AUC), on 100 test data sets consisting of a random subset of samples (80%) to predict case status in *Klebsiella* colonized patients (N = 232) using different taxonomical data inputs. Black circles indicate median AUC values, and black lines indicate standard deviation (*p* indicates Tukey multiple pairwise-comparison ANOVA *p-*value following one way). (B) Top model features for regularized logistic regression models using amplicon sequence variants (ASVs) as input data, corresponding to panel A, “ASVs.” Circles indicate mean feature importance and lines indicate interquartile range. Feature importance values in red and black indicate a regression weight that are weighted toward cases and controls, respectively. (C) Regularized logistic regression model performance on test data sets for 100 seeds predicting case status in *Klebsiella* colonized patients using all ASVs (All ASVs) or excluding ASVs ASV000001, ASV000021, and ASV000195 (-Top ASVs, *p* indicates student’s *t* test *p-*value). Black circles indicate median AUC values, and black lines indicate standard deviation. For panels A and C, each datapoint indicates one test data set.

Consistent with previous observations that gut dominance by *Klebsiella* is a risk factor for infection in colonized patients (8–10), ASV000001 *Klebsiella* was the most important feature in our regularized logistic regression models and was weighted toward cases (Figure 2B). Interestingly, two other ASVs, ASV000021 *Akkermansia*, and ASV000195 *Anaerostipes*, were also highly important features weighted toward cases (Figure 2B). This suggests that other members of the gut microbiota have discriminatory power for case status, rather than discriminatory power being limited to *Klebsiella*. Similar results were yielded in our random forest models (Figure S2A). Given the relatively high feature importance of these ASVs compared to other important features (Figure 2B), we hypothesized that removal of the ASVs may result in a model with no ability to classify cases and controls (AUC ≤ 0.5). Removal of these ASVs significantly reduced model performance (Figure 2C); however, most models were still able to classify cases and controls better than chance (AUC > 0.5). This indicates that peak model performance relies on inclusion of many or all ASVs, rather than a small subset of ASVs.

### Case and control gut community profiles differ

Given that cases and controls can be distinguished based on ASVs using machine learning, we next wanted to determine if the gut community profile of cases and controls differ. To this end, Yue and Clayton θ dissimilarity index was calculated for each patient and used to assess the difference in beta-diversity between cases and controls. Visualization of distances using principal coordinates analysis revealed subtly different clustering of these groups (Figure 3A). Though variance between the two groups was highly dimensional, as indicated by the low axis loadings (Figure 3A), the gut microbiota of cases and controls was significantly different (adjusted *p*-value = 1 x 10^-4^, AMOVA). Only community evenness (Shannon) was significantly different between cases and controls, though community richness and diversity displayed similar trends (Figure 3B-D). Interestingly, the ASVs that were highly important for classifying cases and controls using machine learning models (Figures 2B, S2B) partially differed from those enriched in either cases or controls. Linear discriminant analysis revealed that, as expected, ASV000001 *Klebsiella* was significantly enriched in cases, though unlike what was observed in the machine learning models, ASV000002 *Enterococcus* was also enriched in cases and ASV000012 *Streptococcus* was enriched in controls (Figure 3E-F). Similar results were yielded using OTUs instead of ASVs to differentiate cases and controls (Figure S3). Network analysis revealed that the gut community of controls was more connected than the gut community of cases (Figure S4), suggesting a more stable gut community. Collectively, these data indicate that significant differences, not limited to *Klebsiella* relative abundance, exist between cases and controls that underpin the ability to discriminate these two groups based on gut community profile.

**Figure 3.**
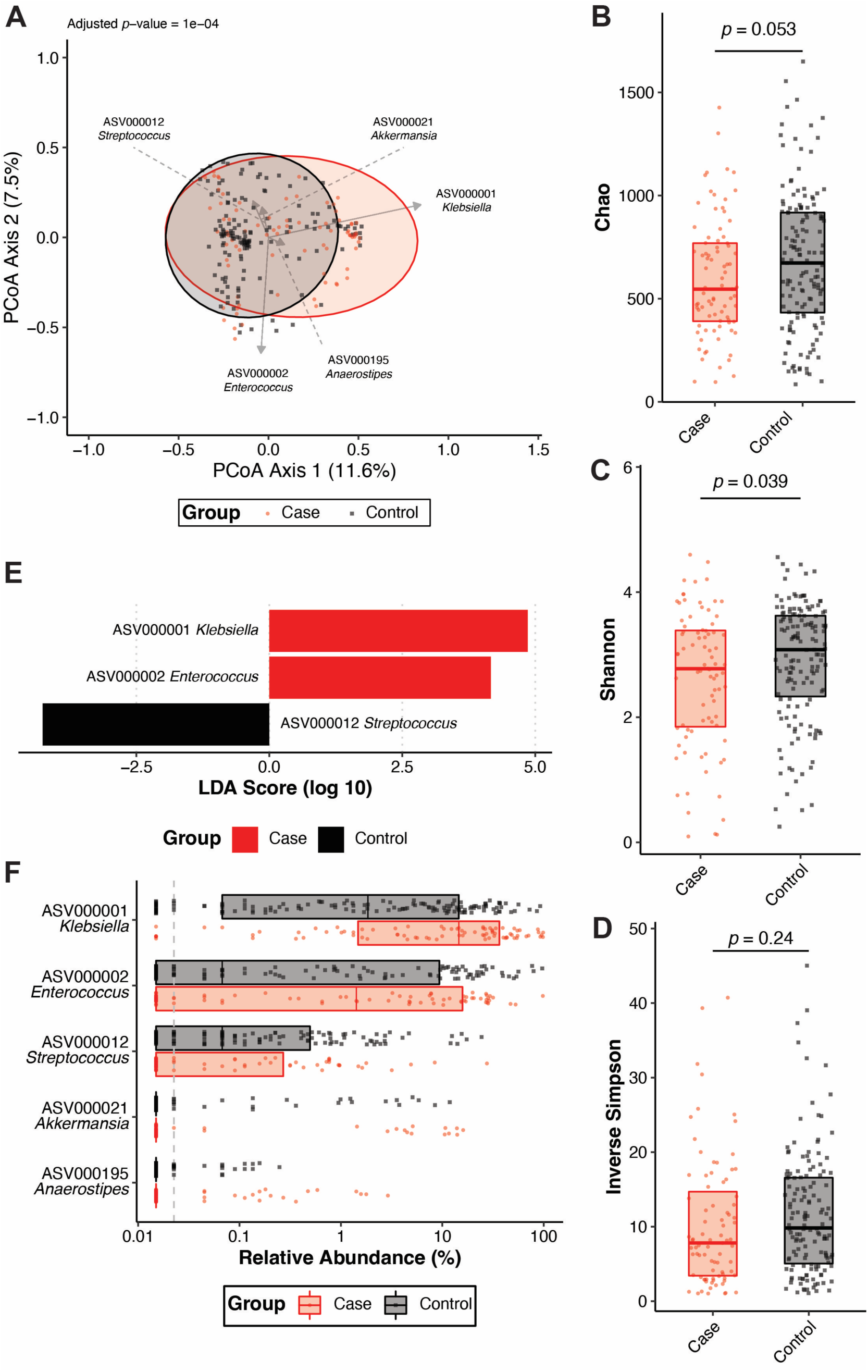
Cases and controls have distinct gut community profiles based on ASVs. (A) Principal coordinates analysis with overlayed biplots of specific ASVs. Analysis of molecular variance (AMOVA) based on the Yue and Clayton θ dissimilarity index was used to assess the difference in beta-diversity between cases (N = 83) and controls (N = 149). Analysis of the (B) Chao, (C) Shannon, and (D) Inverse Simpson alpha-diversity indices between cases (N = 83) and controls (N = 149, boxplot indicates median with interquartile range, *p* indicates student’s *t* test *p-*value). (E) Linear discriminant analysis (LDA) effect size was used to identify differentially abundant (*p-*value < 0.05) ASVs between cases (N = 83) and controls (N = 149). (F) Summary of relative abundances of ASVs that were differentially abundant (Figure 3E) between cases (N = 83) and controls (N = 149) or highly important features for classification of cases and controls using regularized logistic regression shown in Figure 2B (boxplot indicates median with interquartile range). For all panels, each datapoint indicates one patient.

Previously, we were able to detect the presence of multiple *Klebsiella* strains in colonized patients (18). A deeper exploration of ASVs revealed 30 ASVs that were classified as *Klebsiella*, and another 10,470 ASVs that were only classified to the level of Enterobacteriaceae. The majority (83.1%, 193/232) of patients had only one detectable *Klebsiella* ASV; however, 9.9% (23/323) of patients had multiple *Klebsiella* ASVs and 6.9% (16/232) had no *Klebsiella* ASVs (Figure S5A) despite microbiological confirmation of *Klebsiella* colonization. Interestingly, we detected ASV000019 *Klebsiella* in several controls, though no cases (Figure S5B). Though ASVs based on the V4 region of the 16S rRNA gene do not provide high-confidence species-level resolution, it was notable that the ASV000019 sequence primarily aligned to members of the *K. oxytoca* complex (22), whereas the ASV000001 sequence primarily aligned to members of the *K. pneumoniae* complex (Table S2). Interestingly, ASV000001 is absent from patients colonized by ASV000019 (Figure S5C). Collectively, these results suggest that strain-level measurement of cocolonization may be possible through targeted genomic sequencing to understand colonization dynamics. More sophisticated sequencing techniques will need to be developed to assay colonization dynamics using discarded rectal swabs.

### Inclusion of gut microbiota data enhances discrimination of cases and controls

Finally, we hypothesized that inclusion of 16S rRNA gene sequencing data with clinical factors and *Klebsiella* genotype would enhance the ability of machine learning models to discriminate cases and controls. To test this hypothesis, we permutated ASVs with patient factors and *Klebsiella* genotype in our regularized logistic regression models. 84 clinical factors, including several laboratory values, antibiotic exposure, and comorbidities were included (Table S3) and the 27 infection-associated genes identified in our previous comparative genomics study were included as *Klebsiella* genotype (18). Clinical data were missing for two patients, so these patients were excluded from all analyses. Use of clinical factors as the sole input variables led to poor model performance (Figure 4): 14/100 of regularized logistic regression models had an AUC ≤ 0.5, with a median AUC = 0.6. Addition of ASVs to clinical factors enhanced median model performance (Figure 4, median AUC = 0.64). The lack of predictive ability of the clinical factors, especially antimicrobial exposure is somewhat surprising, as gut dominance is a known risk factor for infection (8–10), and disruption of the gut microbiota, such as what occurs with antibiotic exposure, leads to dominance in experimental gut colonization models (11, 13). Therefore, one may expect that antibiotic exposure would be an important feature for discriminating cases and controls in this study. Rather, exposure to most antibiotics was not amongst the most important features in regularized logistic regression models using clinical factors as the input variables (Figure S6A) and the effects of antibiotic exposure on model performance was negligible (Figure S6B). This included a variable for “high-risk” antibiotic exposure, which is a composite variable that includes β-lactam/β-lactamase inhibitor combinations, carbapenems, third- and fourth-generation cephalosporins, fluoroquinolones, clindamycin, and oral vancomycin based on their impact on indigenous gut microbiota (23). The only antibiotic present amongst the most important features was aminoglycoside exposure, and its effects on model performance was subtle (Figure S6A). The importance of antibiotics was further reduced when ASVs were included (Figure S6C-D).

**Figure 4.**
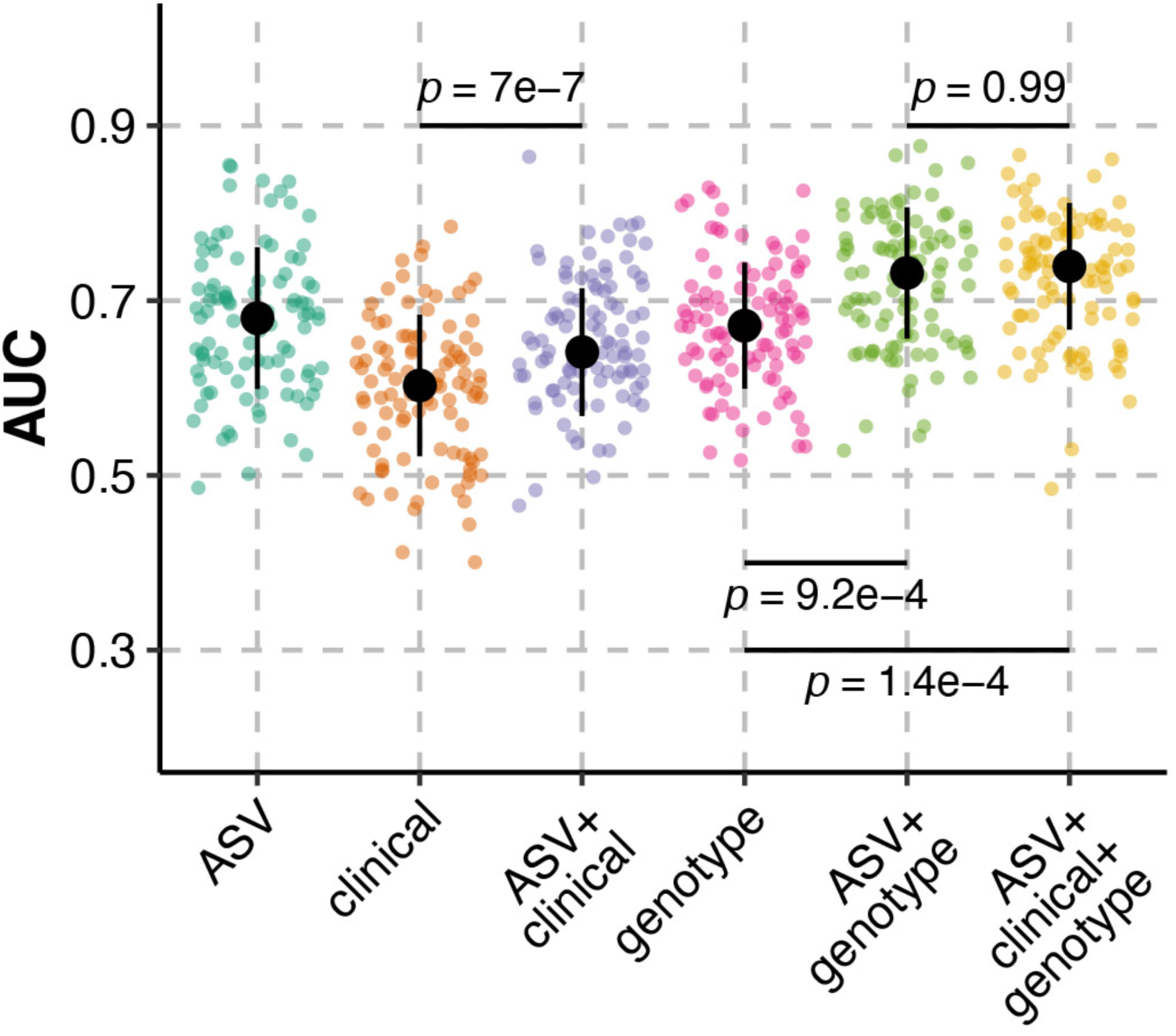
Inclusion of ASVs enhances the ability to discriminate cases from controls. Regularized logistic regression model performance, as measured by area under the receiver-operator characteristic curve (AUC), on test data sets for 100 seeds predicting case status in *Klebsiella* colonized patients (N = 230) using combinations of clinical variables, *Klebsiella* genotype, and ASVs. Black circles indicate median AUC values, black lines indicate standard deviation, and *p* indicates Tukey multiple pairwise-comparison ANOVA *p-*value following one way.

Use of *Klebsiella* genotype as the sole input variables led to a median model performance that was greater than that of clinical factors alone (Figure 4, median AUC = 0.67). This finding is expected, as the 27 genes used as input variables are known to be associated with cases in our previous study (18), whereas most clinical factors were not associated with case status in our original cohort study (7). Interestingly, addition of ASVs to *Klebsiella* genotype enhanced model performance, leading to a median model performance exceeding 0.7 (Figure 4, median AUC = 0.73). Finally, integration of all three datasets led to the highest median model performance, though performance was similar to models using only ASVs and *Klebsiella* genotype (Figure 4, median AUC = 0.74). Similar results were yielded using random forest; however, inclusion of clinical factors with ASVs and *Klebsiella* genotype reduced median model performance (Figure S7, median AUC = 0.74) compared to only ASVs and *Klebsiella* genotype (Figure S7, median AUC = 0.77). In total, these data indicate that inclusion of ASVs with *Klebsiella* genotype leads to peak model performance. This suggests that the gut community profile patients colonized by *Klebsiella* can be combined with other infection-associated variables to discriminate, and potentially predict, infection in these patients with reasonable confidence.

## Discussion

In this study, we have described the gut community of *Klebsiella*-colonized patients and demonstrated that the gut community differs between patients that remain asymptomatic (controls) and those that acquire a subsequent symptomatic infection with their colonizing strain (cases). In machine learning models based on this data, *Klebsiella* relative abundance had the greatest feature importance other gut microbes were also informative. Interestingly, clinical factors such as antibiotic exposure poorly discriminated cases and controls, whereas a combination of gut community data and *Klebsiella* genotype classified cases and controls with a reasonable degree of accuracy (AUC > 0.7). Collectively, this study demonstrates that the gut community of *Klebsiella*-colonized patients can be integrated with other biomarkers to assess infection risk.

An important facet of this study population compared to other study populations is the diversity of colonizing *Klebsiella* strains. Often, studies aimed at describing the gut microbiota of *Klebsiella*-colonized patients capture patients colonized with highly clonal multi-drug resistant (MDR) *Klebsiella* strains (24, 25). In contrast, >100 unique sequence types of *Klebsiella* were identified in this study population, predominantly from non-MDR lineages (7). The attention given to MDR lineages is of course warranted; however, the majority of *Klebsiella* infections are caused by non-MDR lineages (26) and studies have demonstrated that the bulk of colonizing *Klebsiella* strains are diverse (27). It is likely that the gut microbiota, clinical, and genetic factors that increase infection risk in patients colonized by MDR *Klebsiella* differ from those colonized by non-MDR *Klebsiella*. Additionally, the *Klebsiella* in the study population was predominantly non-hypervirulent strains (e.g., “classical” *Klebsiella*). It is also likely that the gut microbiota, clinical, and genetic factors differ in individuals colonized by hypervirulent *Klebsiella* differ from those colonized by non-hypervirulent *Klebsiella*. For example, we identified a *Klebsiella* factor canonically associated with hypervirulence, the *ter* operon, as a microbiome-dependent gut fitness factor (11). This locus was associated with infection in a hospital-wide patient cohort (28) but not in this cohort of intensive care and hematology/oncology patients (7). This highlights the importance of studying all lineages with pathogenic potential to enable accurate risk assessment in colonized patients to reduce the burden of *Klebsiella* disease.

Many microbiome studies classify individuals at risk for or experiencing disease as being in a state of dysbiosis; however, this imprecise term often lacks the context of the definition of a healthy microbiome. This is critical for establishing a causal link between the gut microbiome and disease, especially as the microbiome gradually shifts with age, environment, diet, healthcare exposure, and yet undiscovered variables (reviewed in (29)). The goal of the present study is not to indicate that the gut microbiome of *Klebsiella*-colonized patients is in a state of health or dysbiosis. Rather, the goal is to identify biomarkers that predict infection in colonized patients. Ideally, the observations here will be tested experimentally to explore a causal role in disease. For example, *Akkermansia* (ASV000021) is currently being considered as a probiotic therapy due to its positive impacts on health (30–32). Yet, in this study, *Akkermansia* is important for model performance (Figure 2B) but is relatively low abundance and not enriched in cases (Figure 3C-D). This finding highlights differences between machine leaning and classic linear discriminant analysis approaches for identifying sequences associated with specific communities. It may be the case that the ASVs identified through linear discriminant analysis have occult interactions with one another and/or other ASVs that explain the differential outcomes of these approaches. Similarly, laboratory experiments have demonstrated that members of the *K. oxytoca* complex can reduce *K. pneumoniae* gut colonization (15). Here we observed that the ASV that most likely represents the majority of the *K. pneumoniae* complex (ASV000001) is absent in patients colonized by the ASV that most likely represents the majority of the *K. oxytoca* complex (ASV000019, Figure S5C). Despite a potential probiotic effect against *K. pneumoniae*, *K. oxytoca* is a pathogen that is often highly antimicrobial resistant (reviewed in (22)). Therefore, while microbial competition with *K. pneumoniae* may explain this finding, characterizing *K. oxytoca* as a member of a healthy or dysbiotic gut microbiome remains in question. Further exploration of the biomarkers identified in this study is necessary to determine their importance in influencing infection risk in *Klebsiella*-colonized patients and therein define dysbiosis and its role in infection risk in this patient population.

The variables that are most important in classifying cases and controls likely differ between pathogens and patient populations. For example, clinical biomarkers do not appear to be critical for discriminating case status in this study (Figure 4). This is in contrast to studies performed at the same clinical site leveraging electronic health records to stratify the risk of complicated *Clostridium* (*Clostridioides*) *difficile* infection (33), suggesting a disease-specific effect where the utility of these clinical data in making predictions varies across prediction tasks. Similarly, the finding that ASVs yield the optimal taxonomical resolution for classifying case status (Figure 2) is interesting. A recent machine learning study determined that OTUs were the optimal taxonomical level for predicting colorectal cancer (34). The preference for use of ASVs or OTUs in microbiome studies remains contested (35, 36); however, our study supports the premise that optimal taxonomical resolution is highly dependent on the patient population and outcomes of interest and does not necessarily favor OTUs or ASVs. Ideally, clinical studies interrogating the role of the microbiome in disease would report both OTU and ASV data when using 16S rRNA gene sequencing, such that we gain a greater understanding of how taxonomical resolution influences the stratification of patient risk in conjunction with other potential risk factors.

Though this study adds to our understanding of the gut microbiome of *Klebsiella*-colonized patients, it is not without its limitations. First, we used a case-control design for this study to carefully control for the influence of known and unknown patient factors. However, this study design leads to an overrepresentation of infection in the study population and the predictive modeling metrics should be interpreted only in the context of this study, since in the general population we would expect a much lower infection risk, such as the 4.3% attack rate in our large cohort study from which this nested case-control study was derived (7). Ideally, future studies assessing the role of the microbiome as a risk factor for *Klebsiella* infection will accurately represent the true attack rate while capturing a large enough number of patients, both colonized by *Klebsiella* and not, to maintain suitable study power. Therefore, hypotheses generated in small- and medium-sized studies can be rigorously tested in a study population that reflects the general population. Second, this study is limited in its ability to make functional conclusions about the microbiome due to the use of 16S rRNA gene sequencing instead of metagenomics or other -omics approaches. Unfortunately, many -omics approaches remain cost restrictive and lack easily testable hypotheses. This and similar studies will aid in the generation of hypotheses that can be tested using these approaches in the future. Finally, the use of machine learning models in this study is a useful means of determining the discriminatory ability of a large set of variables but is limited in its interpretability. Clinically actionable risk stratification models should be comprised of a small set of easily observable variables. In our previous studies, we developed practical tools for identifying biomarkers in *Klebsiella*-colonized patients including measurement of *Klebsiella* relative abundance and detection of infection-associated genes by PCR (7, 8). We hope that additional practical tools to assess the role of the microbiome in infection risk in *Klebsiella*-colonized patients will be developed and integrated with our previously developed tools.

The addition of this study to our collection of studies assessing patient factors, gut dominance, and *Klebsiella* genotype (7, 8, 18) represents one of the most comprehensive explorations of infection risk in a cohort of *Klebsiella*-colonized patients. Ultimately, this study provides a foundational framework for the development of integrated, actionable models for predicting and stratifying infection risk in *Klebsiella*-colonized patients.

## Methods

### Ethics statement and study subject selection

Patient enrollment and sample collection at the University of Michigan were approved by and performed per the Institutional Review Boards (IRB) of the University of Michigan Medical School (Study number HUM00123033). This study was performed with a waiver of informed consent since the research involves no more than minimal risk to the subjects, could not practicably be carried out without the waiver, and uses discarded samples. Cohort identification, enrollment, clinical data extraction, chart review, case definitions, and case-control matching criteria are described in detail elsewhere (7, 8, 18). Study subjects were selected based on matching criteria, the availability of rectal swab DNA (8), and whole-genome sequencing data corresponding to the colonizing *Klebsiella* strain (18). All infectious *Klebsiella* isolates were concordant with the colonizing strain in the same patients based on Sanger sequencing of the *wzi* locus (18).

### 16S rRNA gene sequencing and data processing

DNA was previously extracted from patient rectal swabs (8) using the MagAttract PowerMicrobiome DNA/RNA Kit (Qiagen) and an epMotion 5075 liquid handling system. Standard PCRs used 1, 2, or 7 μL of undiluted DNA and touchdown PCR used 7 μL of undiluted DNA to amplify the V4 region of the 16S rRNA gene. Sequencing was performed as previously described (37). 16S rRNA gene sequences were processed with mothur (v. 1.48.0) (19, 38). The sequencing error rate was assessed using a predefined mock community and estimated to be 0.033%. Sequences were aligned to the SILVA reference alignment, release 132 (39) and binned into OTUs using the OptiClust method (40) based on 97% sequence similarity or kept as unique sequences for ASVs. Taxonomic composition was assigned by classifying sequences within mothur using a modified version of the Ribosomal Database Project training set, version 18 (41, 42). Data processing was performed using the Great Lakes High-Performance Computing Cluster at the University of Michigan, Ann Arbor or the Carbonate large-memory computer cluster at Indiana University.

## Data analysis

Data analysis was carried out in RStudio 2021.09.0+351 "Ghost Orchid" Release for macOS or in R, v. 4.2.0. R was used instead of RStudio when the analysis was being performed on The Great Lakes High-Performance Computing Cluster at the University of Michigan, Ann Arbor or the Carbonate large-memory computer cluster at Indiana University. For all analyses except network analysis, sample read counts were rarefied to the lowest-abundance sample (4,438 reads). Alpha- and beta-diversity, principal coordinates analysis, and community typing were performed using mothur. θ_YC_ was used as the distance metric for principal coordinates analysis. Differences in community structure were assessed by AMOVA from the vegan package, v. 2.6-2 (43). Differences in alpha-diversity indices were assessed by student’s *t*-test using the stats package, v. 3.6.2. Assessment of differentially enriched OTUs and ASVs was performed with linear discriminant analysis effect size analysis. Supervised machine learning was performed using mikropml, v. 1.4.0 (44). First, continuous data were split into quartiles, then input data was preprocessed in mikropml using the default settings. Supervised machine learning was performed using case status as the outcome. Input data was split 80:20 into train and test groups. An optimal model was trained using 100X 5-fold cross-validation and model performance was evaluated using the test data. For regularized logistic regression, hyperparameter selection was semi-automated. Each model was trained with alpha values ranging from 0 to 1, iterated in steps of 0.1, permutated with lambda values ranging from 10^-4^ to 10^1^, iterated in steps of 3 between each log (e.g., 10^-4^, 2.5x10^-4^, 5x10^-4^, 7.5x10-4, 10^-3^, 2.5x10^-3^…10^1^). Trained model performance was assessed by area under the receiver-operator characteristic curve, and hyperparameters that yielded the best performance were selected to evaluate model performance using the test data. For random forest models, the default hyperparameters were used. For each method, this process was parallelized 100 times, using 100 different seeds to determine the train:test data split, and feature importance and weight (only regularized logistic regression) was determined for all variables. Network analysis was performed using NetCoMi v. 1.1.0 (45). Networks were constructed using the compositionally aware correlation estimators, SparCC (46), and networks were compared by permutation test with 100 permutations. For all analyses, a *p*-value ≤ 0.05 after multiple comparison adjustment was considered statistically significant. Data were visualized using ggplot2, v.4.1.2 (47).

## Data and code availability

The sequencing data generated in this study have been deposited in the Sequence Read Archive (SRA) database under accession PRJNA789565. Deidentified human data are available under restricted access and can be obtained from MAB within 1 year upon request, pending approval from the University of Michigan Institutional Review Board. All other source data and code are available at https://github.com/jayvorn.

## Acknowledgements

The authors would like to thank the University of Michigan Microbiome Core for their assistance with 16S rRNA gene sequencing and to Dr. Anna Seekatz for her valuable insight into all things gut microbiome. This research was supported in part through computational resources and services provided by Advanced Research Computing (ARC), a division of Information and Technology Services (ITS) at the University of Michigan, Ann Arbor. The authors acknowledge the Indiana University Pervasive Technology Institute for providing supercomputing and storage resources that have contributed to the research results reported within this paper.

## Contributions

Conceptualization: JV, KR, MAB

Methodology: JV, KR

Investigation: JV

Visualization: JV

Funding acquisition: JV, MAB

Project administration: MAB

Supervision: MAB

Writing – original draft: JV

Writing – review & editing: JV, KR, MAB

## Supplementary Information

**Figure S1.**
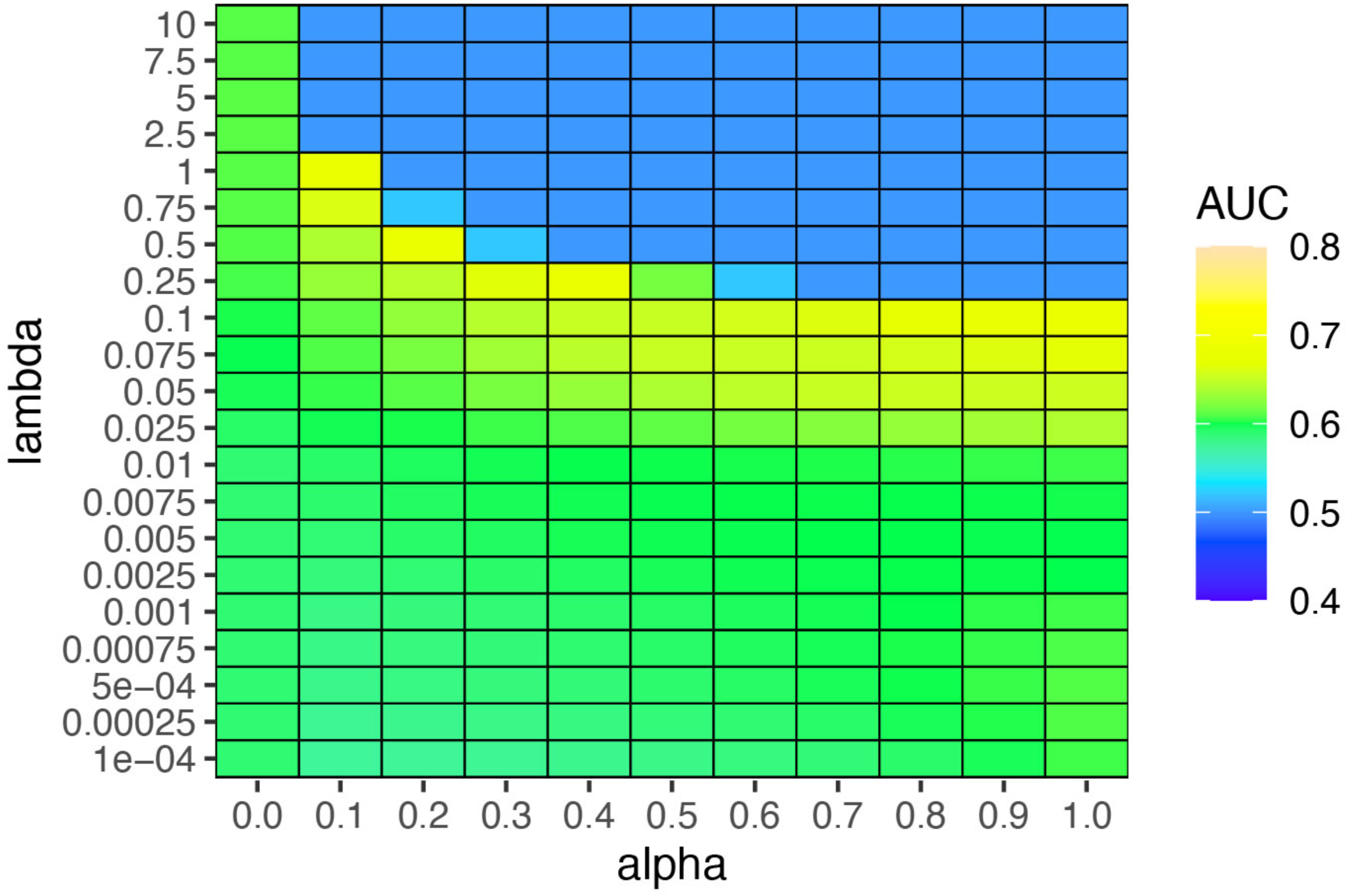
Example of regularized logistic regression hyperparameter selection. Regularized logistic regression trained model performance as measured by area under the receiver-operator characteristic curve (AUC) for 100 seeds. ASVs were used as input data. Each box contains the mean AUC values for each hyperparameter combination. Future model testing was performed using the hyperparameter combination that yielded the peak AUC for a given seed.

**Figure S2.**
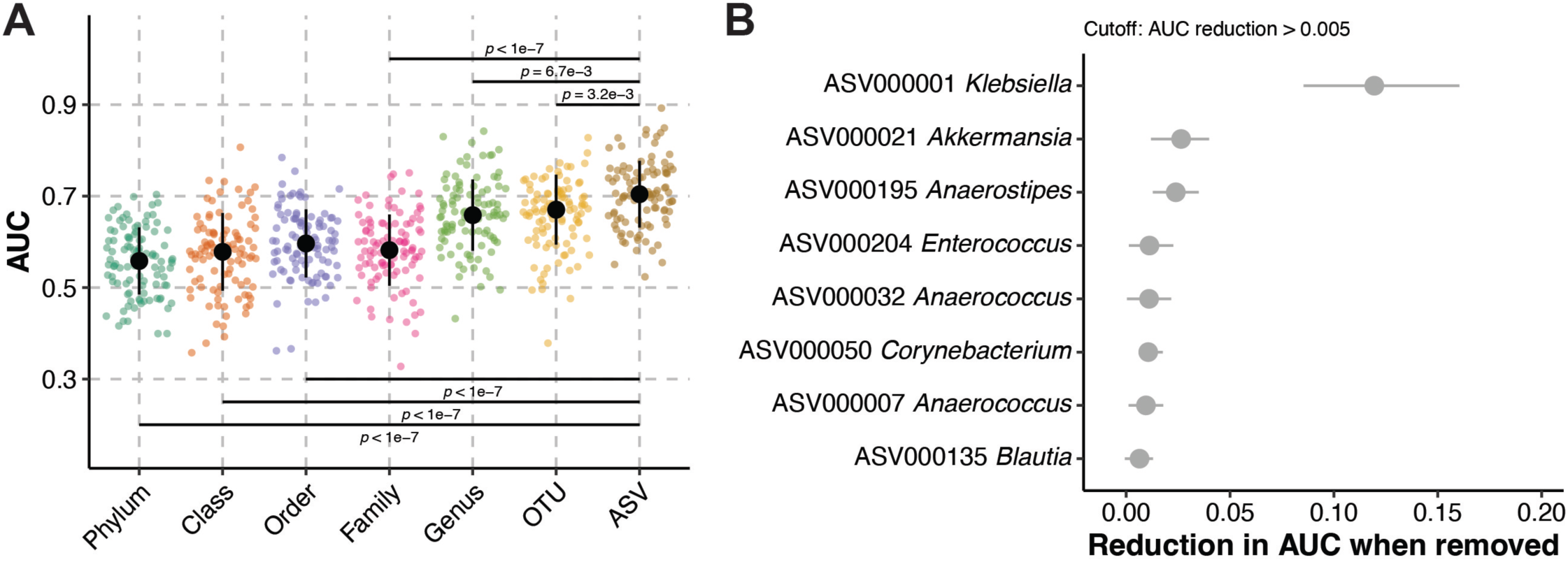
Random forest model performance. (A) Random forest model performance, as measured by area under the receiver-operator characteristic curve (AUC), on test data sets for 100 seeds predicting case status in *Klebsiella* colonized patients (N = 232). Black circles indicate median AUC values, black lines indicate standard deviation, and *p* indicates Tukey multiple pairwise-comparison ANOVA *p-*value following one way. (B) Top model features using amplicon sequence variants (ASVs) as input data, corresponding to panel A, “ASVs.” Circles indicate mean feature importance and lines indicate interquartile range.

**Figure S3.**
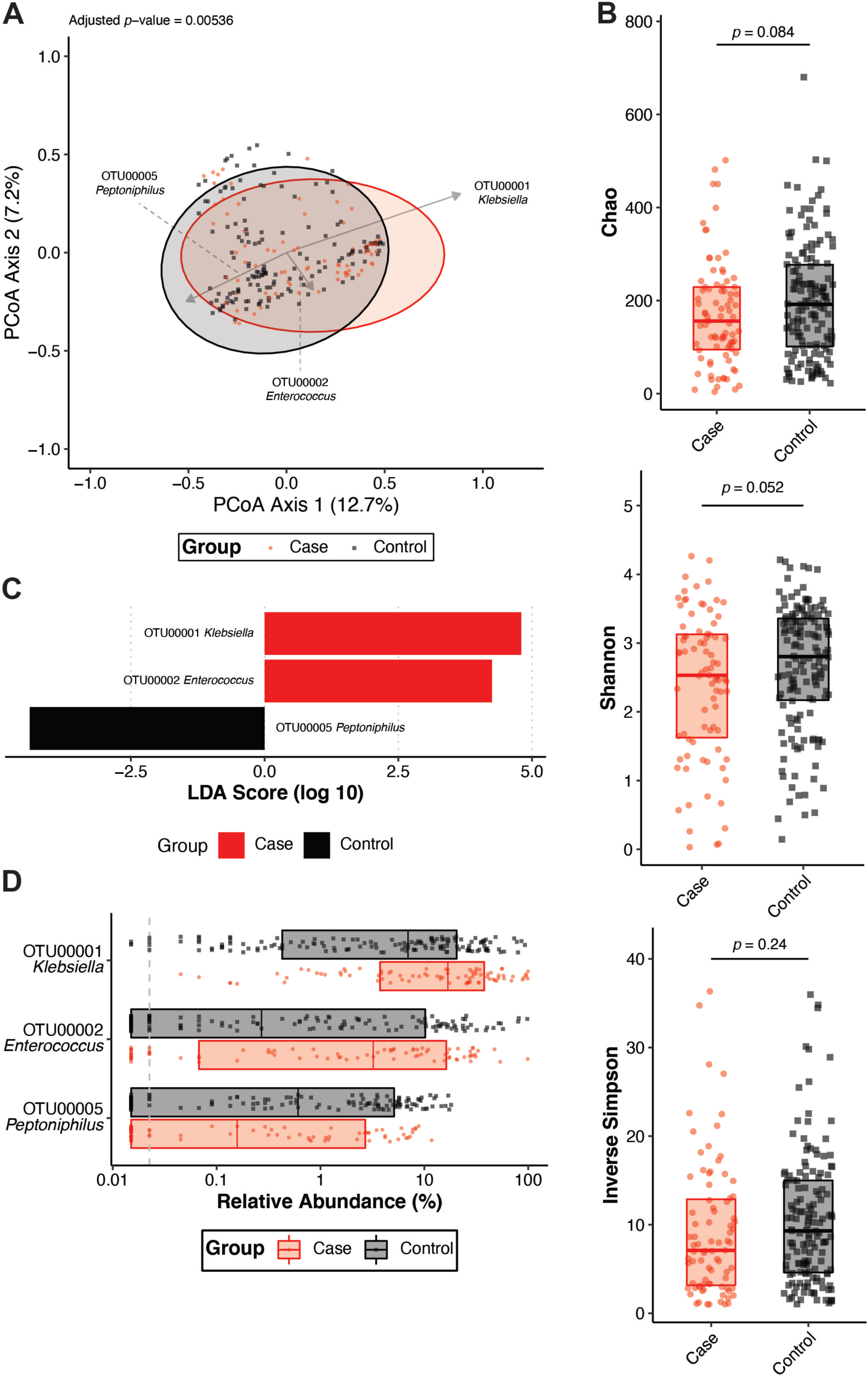
Cases and controls have distinct gut community profiles based on OTUs. (A) Principal coordinates analysis with overlayed biplots of specific OTUs. Analysis of molecular variance (AMOVA) based on the Yue and Clayton θ dissimilarity index was used to assess the difference in beta-diversity between cases (N = 83) and controls (N = 149). (B) Analysis of the Chao, Shannon, and Inverse Simpson alpha-diversity indices between cases (N = 83) and controls (N = 149, boxplot indicates median with interquartile range, *p* indicates student’s *t* test *p-*value). (C) Linear discriminant analysis (LDA) effect size was used to identify differentially abundant OTUs between cases (N = 83) and controls (N = 149). (D) Summary of relative abundances of OTUs that were differentially abundant between cases (N = 83) and controls (N = 149, boxplot indicates median with interquartile range). For all panels, each datapoint indicates one patient.

**Figure S4.**
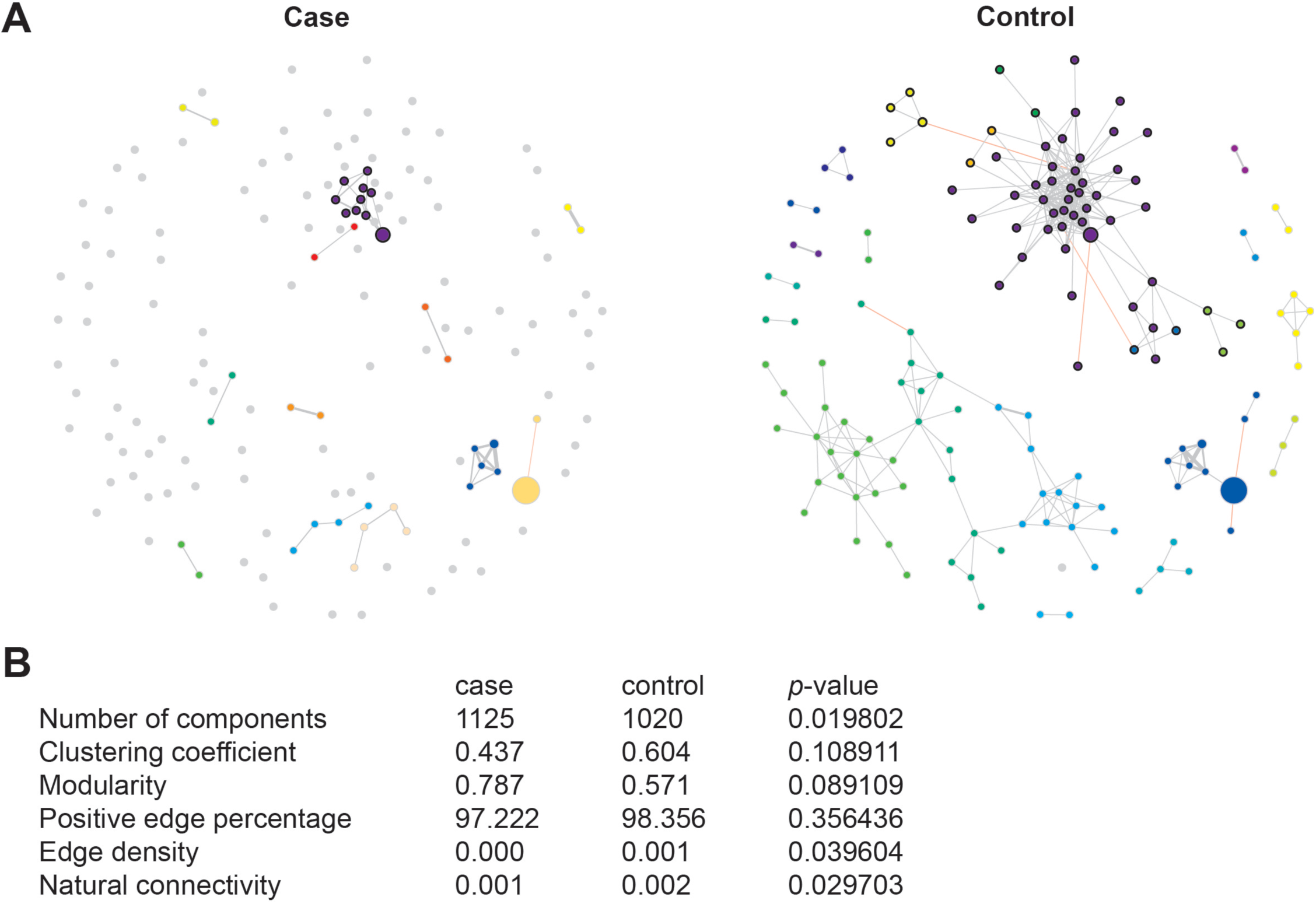
Cases and controls have distinct gut community networks. (A) Network plots of case and control gut communities. Only ASVs with 1,000 total reads were included in network construction, and only nodes with significant correlations (student’s *t*-test Benjamini & Hochberg corrected *p-*value < 0.05) in either group are shown. Each node is a single ASV, scaled to the total read count. Only nodes Black edges are positive correlations, and red edges are negative correlations. Node colors indicate distinct clusters. (B) Permutation test (N = 100 permutations) results comparing case and control network properties.

**Figure S5.**
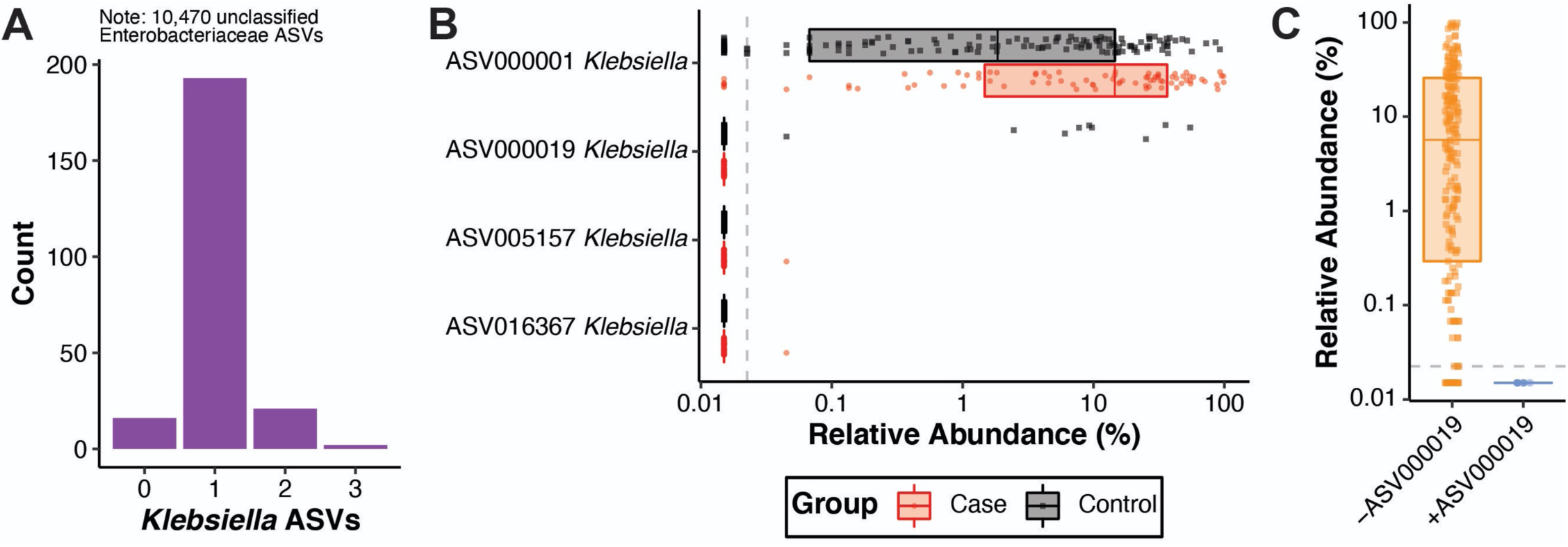
Multiple *Klebsiella* ASVs are present in *Klebsiella*-colonized patients. (A) Count of the number of *Klebsiella* ASVs in each patient (N = 232). (B) Summary of relative abundances of *Klebsiella* ASVs with a sequence count >1 stratified by cases (N = 83) and controls (N = 149, boxplot indicates median with interquartile range). (C) Summary of relative abundance of ASV000001 when ASV000019 is absent (-ASV000019, N = 223) or present (+ASV000019, N = 9, boxplot indicates median with interquartile range). For panels B and C, each datapoint indicates one patient.

**Figure S6.**
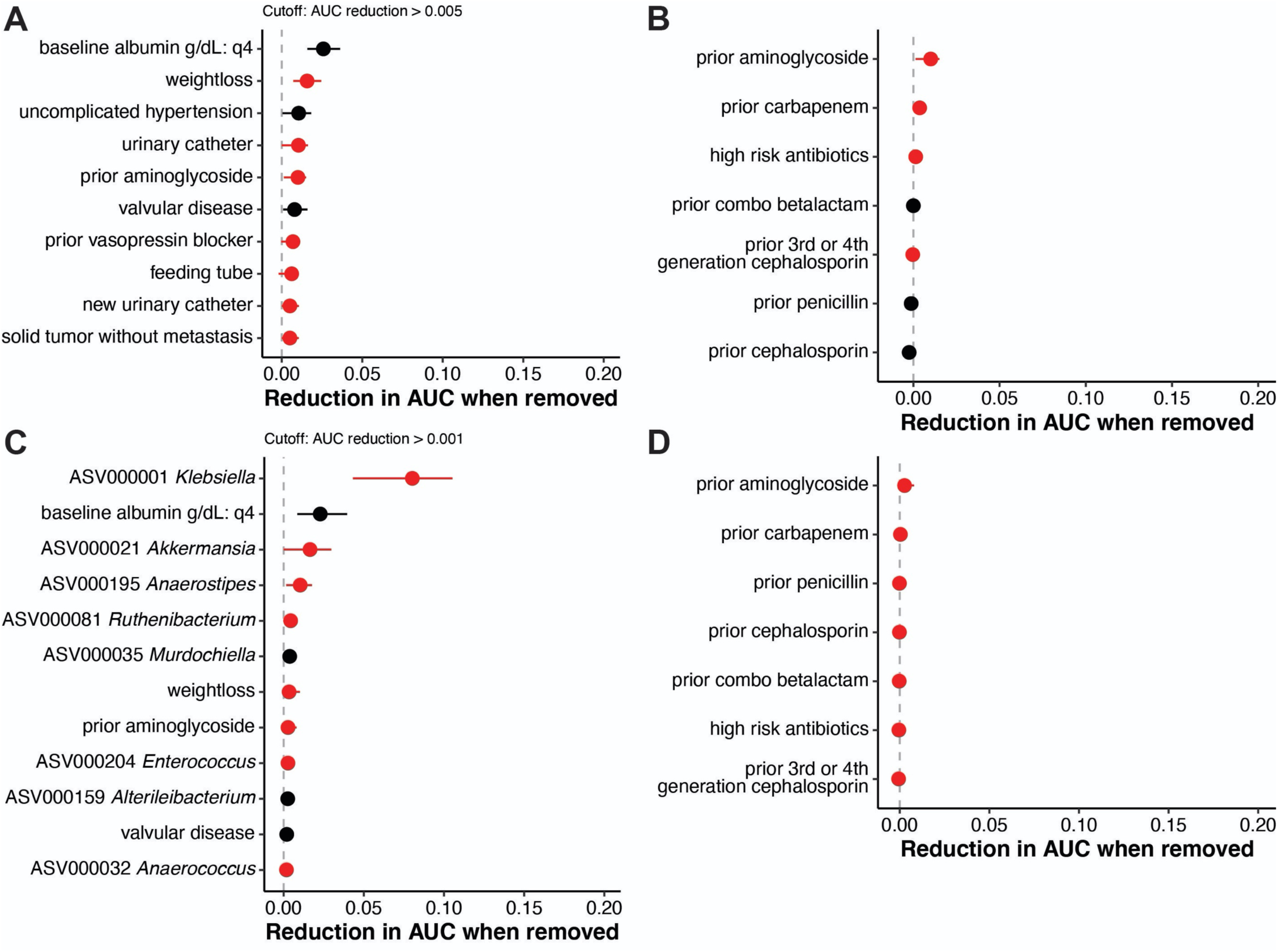
Antibiotic exposure is not important for classifying cases and controls. (A) Top model features for regularized logistic regression models using clinical variables as input data, corresponding to Figure 4 “clinical.” (B) Antibiotic exposure features for regularized logistic regression models using clinical variables as input data, corresponding to Figure 4 “clinical.” (C) Top model features for regularized logistic regression models using clinical variables and ASVs as input data, corresponding to Figure 4 “ASV+clinical.” (D) Antibiotic exposure features for regularized logistic regression models using clinical variables as input data, corresponding to Figure 4 “ASV+clinical.” For all panels, circles indicate mean feature importance and lines indicate interquartile range. Feature importance values in red and black indicate a regression weight that are weighted toward cases and controls, respectively.

**Figure S7.**
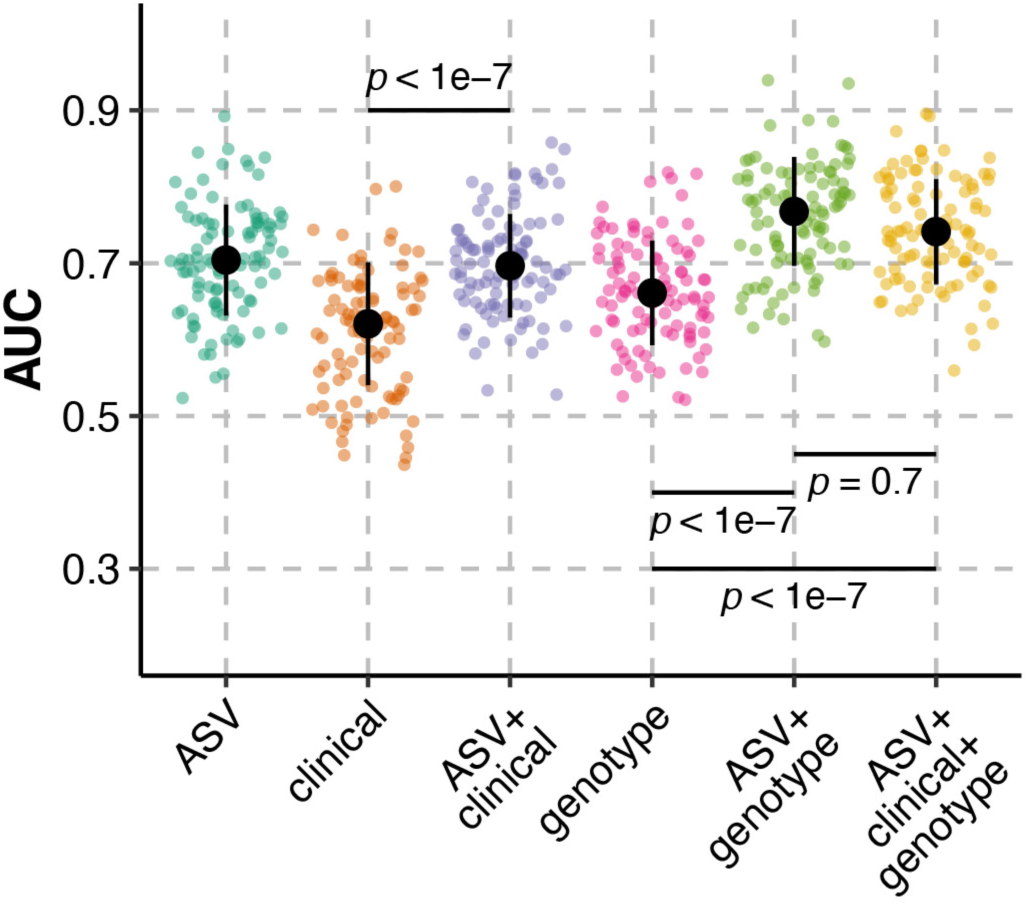
Inclusion of ASVs enhances the ability to discriminate cases from controls using random forest. Random forest model performance, as measured by area under the receiver-operator characteristic curve (AUC), on test data sets for 100 seeds predicting case status in *Klebsiella* colonized patients (N = 230) using various input datasets. Black circles indicate median AUC values, black lines indicate standard deviation, and *p* indicates Tukey multiple pairwise-comparison ANOVA *p-*value following one way.

**Table S1.**
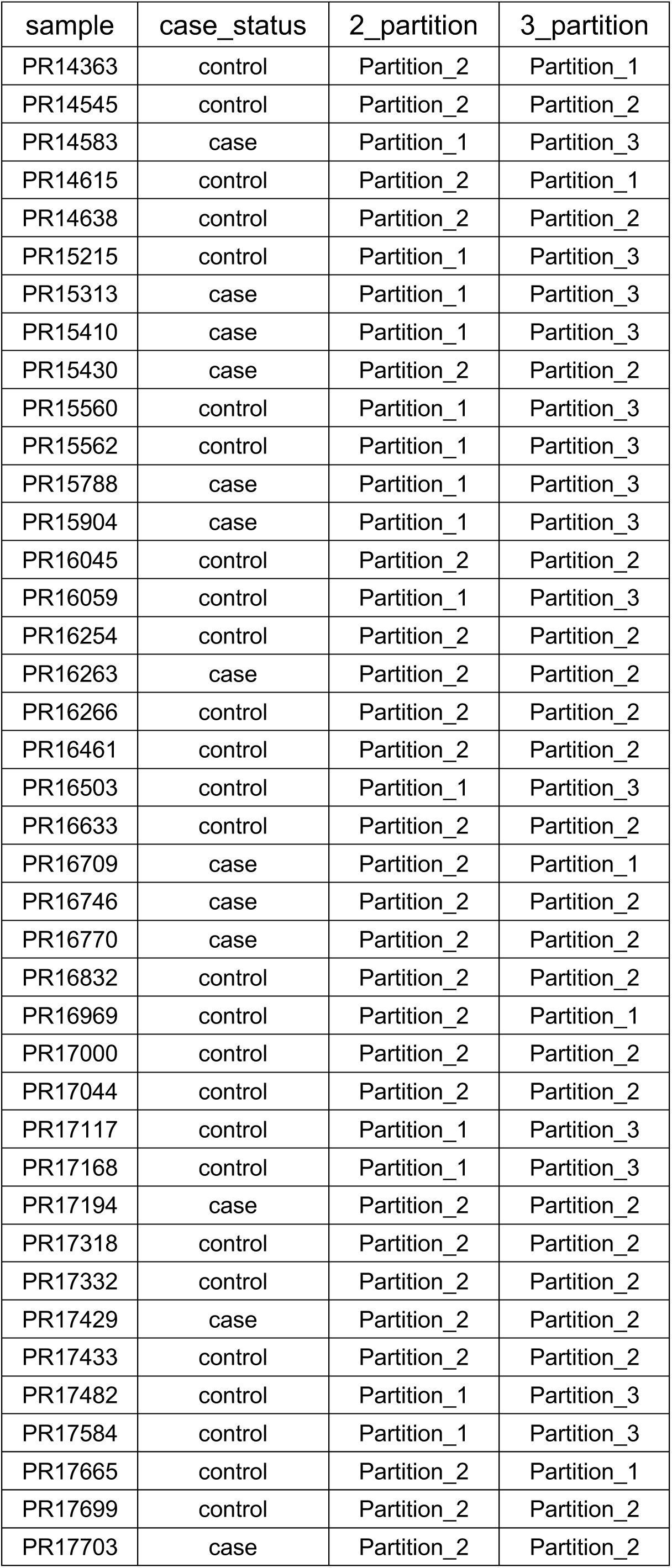
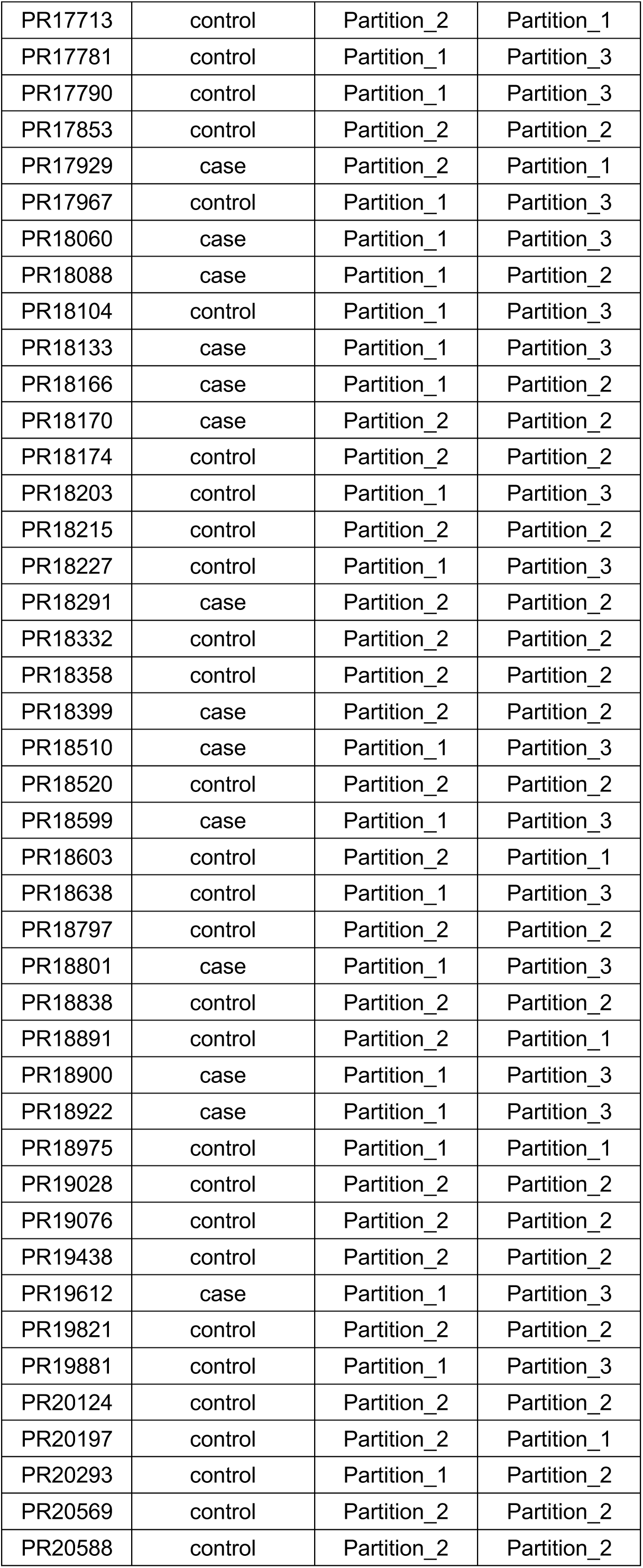
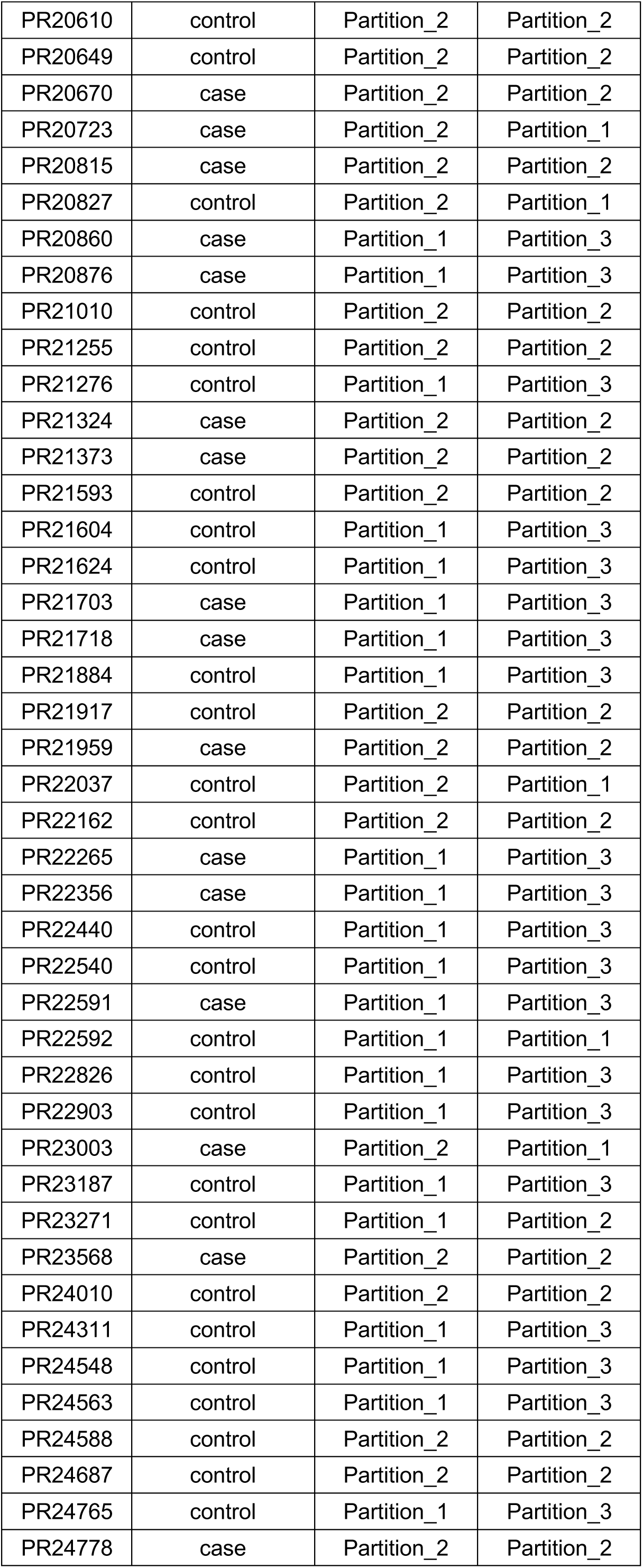
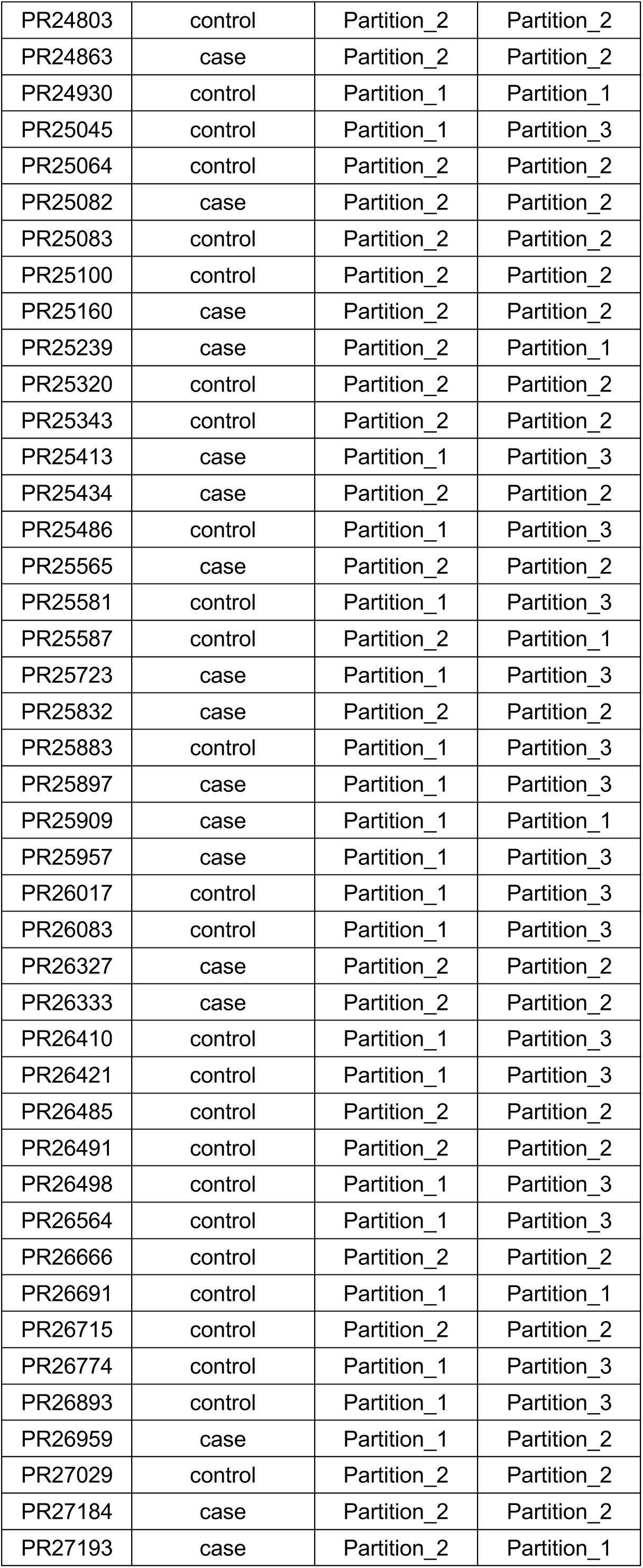
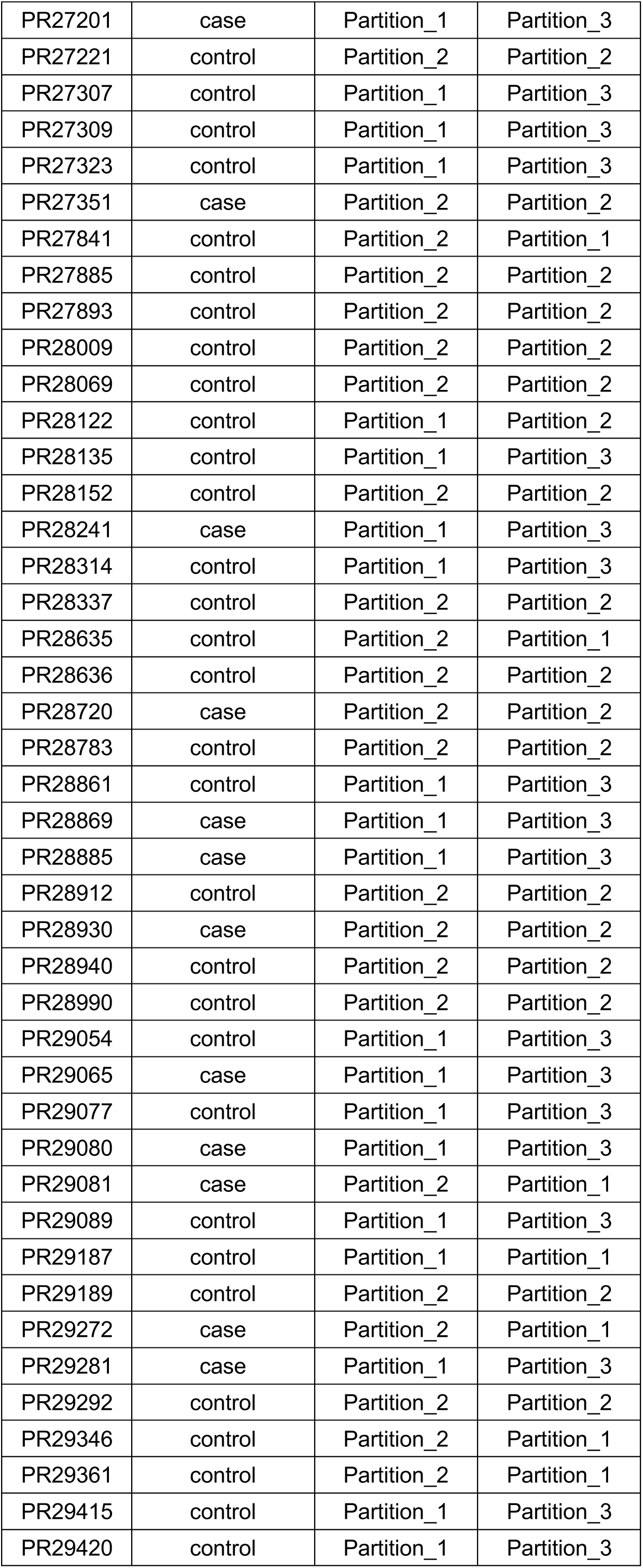
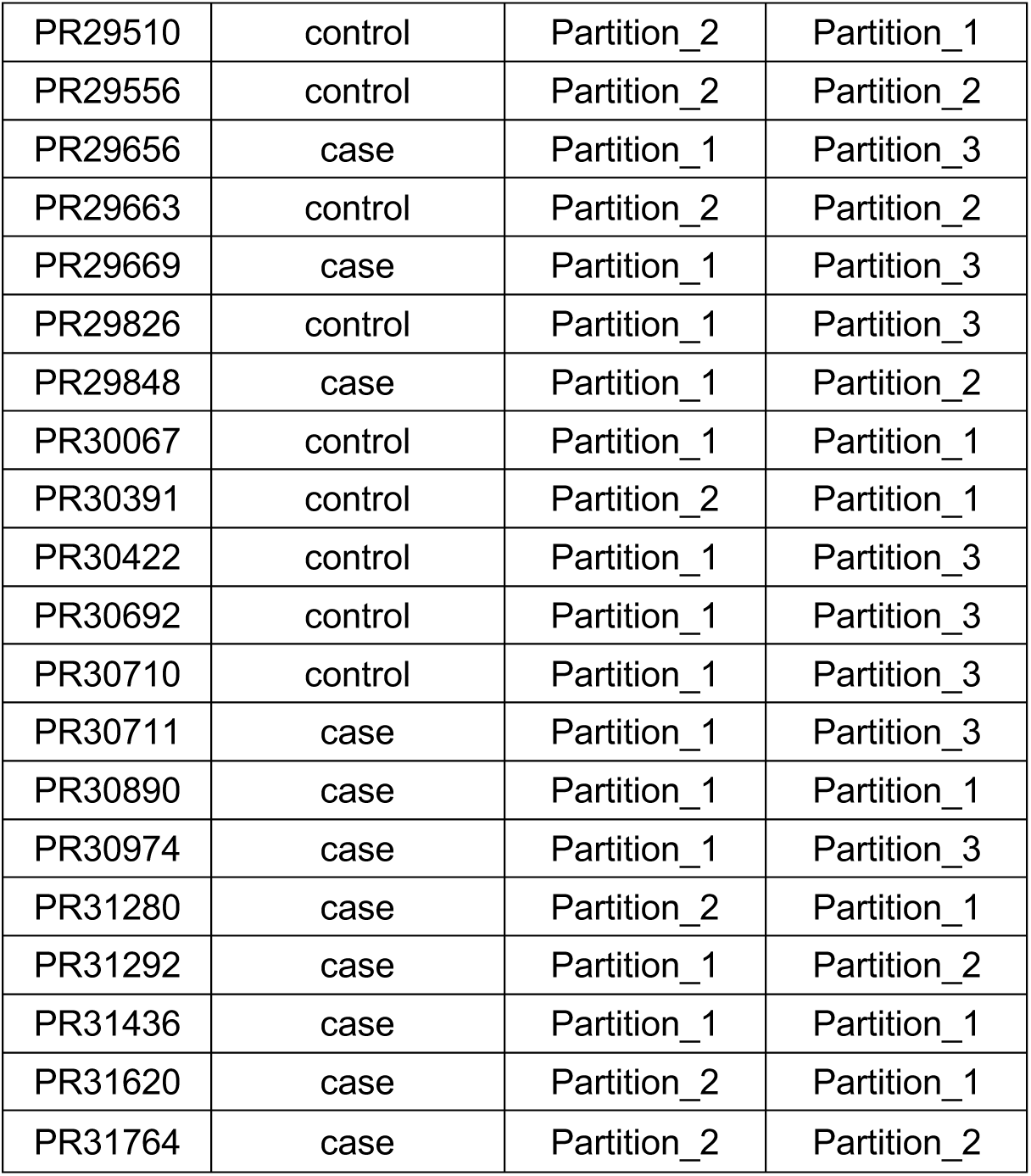
Sample partitions

| sample | case_status | 2_partition | 3_partition |
| --- | --- | --- | --- |
| PR14363 | control | Partition_2 | Partition_1 |
| PR14545 | control | Partition_2 | Partition_2 |
| PR14583 | case | Partition_1 | Partition_3 |
| PR14615 | control | Partition_2 | Partition_1 |
| PR14638 | control | Partition_2 | Partition_2 |
| PR15215 | control | Partition_1 | Partition_3 |
| PR15313 | case | Partition_1 | Partition_3 |
| PR15410 | case | Partition_1 | Partition_3 |
| PR15430 | case | Partition_2 | Partition_2 |
| PR15560 | control | Partition_1 | Partition_3 |
| PR15562 | control | Partition_1 | Partition_3 |
| PR15788 | case | Partition_1 | Partition_3 |
| PR15904 | case | Partition_1 | Partition_3 |
| PR16045 | control | Partition_2 | Partition_2 |
| PR16059 | control | Partition_1 | Partition_3 |
| PR16254 | control | Partition_2 | Partition_2 |
| PR16263 | case | Partition_2 | Partition_2 |
| PR16266 | control | Partition_2 | Partition_2 |
| PR16461 | control | Partition_2 | Partition_2 |
| PR16503 | control | Partition_1 | Partition_3 |
| PR16633 | control | Partition_2 | Partition_2 |
| PR16709 | case | Partition_2 | Partition_1 |
| PR16746 | case | Partition_2 | Partition_2 |
| PR16770 | case | Partition_2 | Partition_2 |
| PR16832 | control | Partition_2 | Partition_2 |
| PR16969 | control | Partition_2 | Partition_1 |
| PR17000 | control | Partition_2 | Partition_2 |
| PR17044 | control | Partition_2 | Partition_2 |
| PR17117 | control | Partition_1 | Partition_3 |
| PR17168 | control | Partition_1 | Partition_3 |
| PR17194 | case | Partition_2 | Partition_2 |
| PR17318 | control | Partition_2 | Partition_2 |
| PR17332 | control | Partition_2 | Partition_2 |
| PR17429 | case | Partition_2 | Partition_2 |
| PR17433 | control | Partition_2 | Partition_2 |
| PR17482 | control | Partition_1 | Partition_3 |
| PR17584 | control | Partition_1 | Partition_3 |
| PR17665 | control | Partition_2 | Partition_1 |
| PR17699 | control | Partition_2 | Partition_2 |
| PR17703 | case | Partition_2 | Partition_2 |
| PR17713 | control | Partition_2 | Partition_1 |
| PR17781 | control | Partition_1 | Partition_3 |
| PR17790 | control | Partition_1 | Partition_3 |
| PR17853 | control | Partition_2 | Partition_2 |
| PR17929 | case | Partition_2 | Partition_1 |
| PR17967 | control | Partition_1 | Partition_3 |
| PR18060 | case | Partition_1 | Partition_3 |
| PR18088 | case | Partition_1 | Partition_2 |
| PR18104 | control | Partition_1 | Partition_3 |
| PR18133 | case | Partition_1 | Partition_3 |
| PR18166 | case | Partition_1 | Partition_2 |
| PR18170 | case | Partition_2 | Partition_2 |
| PR18174 | control | Partition_2 | Partition_2 |
| PR18203 | control | Partition_1 | Partition_3 |
| PR18215 | control | Partition_2 | Partition_2 |
| PR18227 | control | Partition_1 | Partition_3 |
| PR18291 | case | Partition_2 | Partition_2 |
| PR18332 | control | Partition_2 | Partition_2 |
| PR18358 | control | Partition_2 | Partition_2 |
| PR18399 | case | Partition_2 | Partition_2 |
| PR18510 | case | Partition_1 | Partition_3 |
| PR18520 | control | Partition_2 | Partition_2 |
| PR18599 | case | Partition_1 | Partition_3 |
| PR18603 | control | Partition_2 | Partition_1 |
| PR18638 | control | Partition_1 | Partition_3 |
| PR18797 | control | Partition_2 | Partition_2 |
| PR18801 | case | Partition_1 | Partition_3 |
| PR18838 | control | Partition_2 | Partition_2 |
| PR18891 | control | Partition_2 | Partition_1 |
| PR18900 | case | Partition_1 | Partition_3 |
| PR18922 | case | Partition_1 | Partition_3 |
| PR18975 | control | Partition_1 | Partition_1 |
| PR19028 | control | Partition_2 | Partition_2 |
| PR19076 | control | Partition_2 | Partition_2 |
| PR19438 | control | Partition_2 | Partition_2 |
| PR19612 | case | Partition_1 | Partition_3 |
| PR19821 | control | Partition_2 | Partition_2 |
| PR19881 | control | Partition_1 | Partition_3 |
| PR20124 | control | Partition_2 | Partition_2 |
| PR20197 | control | Partition_2 | Partition_1 |
| PR20293 | control | Partition_1 | Partition_2 |
| PR20569 | control | Partition_2 | Partition_2 |
| PR20588 | control | Partition_2 | Partition_2 |
| PR20610 | control | Partition_2 | Partition_2 |
| PR20649 | control | Partition_2 | Partition_2 |
| PR20670 | case | Partition_2 | Partition_2 |
| PR20723 | case | Partition_2 | Partition_1 |
| PR20815 | case | Partition_2 | Partition_2 |
| PR20827 | control | Partition_2 | Partition_1 |
| PR20860 | case | Partition_1 | Partition_3 |
| PR20876 | case | Partition_1 | Partition_3 |
| PR21010 | control | Partition_2 | Partition_2 |
| PR21255 | control | Partition_2 | Partition_2 |
| PR21276 | control | Partition_1 | Partition_3 |
| PR21324 | case | Partition_2 | Partition_2 |
| PR21373 | case | Partition_2 | Partition_2 |
| PR21593 | control | Partition_2 | Partition_2 |
| PR21604 | control | Partition_1 | Partition_3 |
| PR21624 | control | Partition_1 | Partition_3 |
| PR21703 | case | Partition_1 | Partition_3 |
| PR21718 | case | Partition_1 | Partition_3 |
| PR21884 | control | Partition_1 | Partition_3 |
| PR21917 | control | Partition_2 | Partition_2 |
| PR21959 | case | Partition_2 | Partition_2 |
| PR22037 | control | Partition_2 | Partition_1 |
| PR22162 | control | Partition_2 | Partition_2 |
| PR22265 | case | Partition_1 | Partition_3 |
| PR22356 | case | Partition_1 | Partition_3 |
| PR22440 | control | Partition_1 | Partition_3 |
| PR22540 | control | Partition_1 | Partition_3 |
| PR22591 | case | Partition_1 | Partition_3 |
| PR22592 | control | Partition_1 | Partition_1 |
| PR22826 | control | Partition_1 | Partition_3 |
| PR22903 | control | Partition_1 | Partition_3 |
| PR23003 | case | Partition_2 | Partition_1 |
| PR23187 | control | Partition_1 | Partition_3 |
| PR23271 | control | Partition_1 | Partition_2 |
| PR23568 | case | Partition_2 | Partition_2 |
| PR24010 | control | Partition_2 | Partition_2 |
| PR24311 | control | Partition_1 | Partition_3 |
| PR24548 | control | Partition_1 | Partition_3 |
| PR24563 | control | Partition_1 | Partition_3 |
| PR24588 | control | Partition_2 | Partition_2 |
| PR24687 | control | Partition_2 | Partition_2 |
| PR24765 | control | Partition_1 | Partition_3 |
| PR24778 | case | Partition_2 | Partition_2 |
| PR24803 | control | Partition_2 | Partition_2 |
| PR24863 | case | Partition_2 | Partition_2 |
| PR24930 | control | Partition_1 | Partition_1 |
| PR25045 | control | Partition_1 | Partition_3 |
| PR25064 | control | Partition_2 | Partition_2 |
| PR25082 | case | Partition_2 | Partition_2 |
| PR25083 | control | Partition_2 | Partition_2 |
| PR25100 | control | Partition_2 | Partition_2 |
| PR25160 | case | Partition_2 | Partition_2 |
| PR25239 | case | Partition_2 | Partition_1 |
| PR25320 | control | Partition_2 | Partition_2 |
| PR25343 | control | Partition_2 | Partition_2 |
| PR25413 | case | Partition_1 | Partition_3 |
| PR25434 | case | Partition_2 | Partition_2 |
| PR25486 | control | Partition_1 | Partition_3 |
| PR25565 | case | Partition_2 | Partition_2 |
| PR25581 | control | Partition_1 | Partition_3 |
| PR25587 | control | Partition_2 | Partition_1 |
| PR25723 | case | Partition_1 | Partition_3 |
| PR25832 | case | Partition_2 | Partition_2 |
| PR25883 | control | Partition_1 | Partition_3 |
| PR25897 | case | Partition_1 | Partition_3 |
| PR25909 | case | Partition_1 | Partition_1 |
| PR25957 | case | Partition_1 | Partition_3 |
| PR26017 | control | Partition_1 | Partition_3 |
| PR26083 | control | Partition_1 | Partition_3 |
| PR26327 | case | Partition_2 | Partition_2 |
| PR26333 | case | Partition_2 | Partition_2 |
| PR26410 | control | Partition_1 | Partition_3 |
| PR26421 | control | Partition_1 | Partition_3 |
| PR26485 | control | Partition_2 | Partition_2 |
| PR26491 | control | Partition_2 | Partition_2 |
| PR26498 | control | Partition_1 | Partition_3 |
| PR26564 | control | Partition_1 | Partition_3 |
| PR26666 | control | Partition_2 | Partition_2 |
| PR26691 | control | Partition_1 | Partition_1 |
| PR26715 | control | Partition_2 | Partition_2 |
| PR26774 | control | Partition_1 | Partition_3 |
| PR26893 | control | Partition_1 | Partition_3 |
| PR26959 | case | Partition_1 | Partition_2 |
| PR27029 | control | Partition_2 | Partition_2 |
| PR27184 | case | Partition_2 | Partition_2 |
| PR27193 | case | Partition_2 | Partition_1 |
| PR27201 | case | Partition_1 | Partition_3 |
| PR27221 | control | Partition_2 | Partition_2 |
| PR27307 | control | Partition_1 | Partition_3 |
| PR27309 | control | Partition_1 | Partition_3 |
| PR27323 | control | Partition_1 | Partition_3 |
| PR27351 | case | Partition_2 | Partition_2 |
| PR27841 | control | Partition_2 | Partition_1 |
| PR27885 | control | Partition_2 | Partition_2 |
| PR27893 | control | Partition_2 | Partition_2 |
| PR28009 | control | Partition_2 | Partition_2 |
| PR28069 | control | Partition_2 | Partition_2 |
| PR28122 | control | Partition_1 | Partition_2 |
| PR28135 | control | Partition_1 | Partition_3 |
| PR28152 | control | Partition_2 | Partition_2 |
| PR28241 | case | Partition_1 | Partition_3 |
| PR28314 | control | Partition_1 | Partition_3 |
| PR28337 | control | Partition_2 | Partition_2 |
| PR28635 | control | Partition_2 | Partition_1 |
| PR28636 | control | Partition_2 | Partition_2 |
| PR28720 | case | Partition_2 | Partition_2 |
| PR28783 | control | Partition_2 | Partition_2 |
| PR28861 | control | Partition_1 | Partition_3 |
| PR28869 | case | Partition_1 | Partition_3 |
| PR28885 | case | Partition_1 | Partition_3 |
| PR28912 | control | Partition_2 | Partition_2 |
| PR28930 | case | Partition_2 | Partition_2 |
| PR28940 | control | Partition_2 | Partition_2 |
| PR28990 | control | Partition_2 | Partition_2 |
| PR29054 | control | Partition_1 | Partition_3 |
| PR29065 | case | Partition_1 | Partition_3 |
| PR29077 | control | Partition_1 | Partition_3 |
| PR29080 | case | Partition_1 | Partition_3 |
| PR29081 | case | Partition_2 | Partition_1 |
| PR29089 | control | Partition_1 | Partition_3 |
| PR29187 | control | Partition_1 | Partition_1 |
| PR29189 | control | Partition_2 | Partition_2 |
| PR29272 | case | Partition_2 | Partition_1 |
| PR29281 | case | Partition_1 | Partition_3 |
| PR29292 | control | Partition_2 | Partition_2 |
| PR29346 | control | Partition_2 | Partition_1 |
| PR29361 | control | Partition_2 | Partition_1 |
| PR29415 | control | Partition_1 | Partition_3 |
| PR29420 | control | Partition_1 | Partition_3 |
| PR29510 | control | Partition_2 | Partition_1 |
| PR29556 | control | Partition_2 | Partition_2 |
| PR29656 | case | Partition_1 | Partition_3 |
| PR29663 | control | Partition_2 | Partition_2 |
| PR29669 | case | Partition_1 | Partition_3 |
| PR29826 | control | Partition_1 | Partition_3 |
| PR29848 | case | Partition_1 | Partition_2 |
| PR30067 | control | Partition_1 | Partition_1 |
| PR30391 | control | Partition_2 | Partition_1 |
| PR30422 | control | Partition_1 | Partition_3 |
| PR30692 | control | Partition_1 | Partition_3 |
| PR30710 | control | Partition_1 | Partition_3 |
| PR30711 | case | Partition_1 | Partition_3 |
| PR30890 | case | Partition_1 | Partition_1 |
| PR30974 | case | Partition_1 | Partition_3 |
| PR31280 | case | Partition_2 | Partition_1 |
| PR31292 | case | Partition_1 | Partition_2 |
| PR31436 | case | Partition_1 | Partition_1 |
| PR31620 | case | Partition_2 | Partition_1 |
| PR31764 | case | Partition_2 | Partition_2 |

**Table S2.** BLAST alignment* to ASV000001 and ASV000019 rRNA sequences

| Query | Subject | Species complex | Accession |
| --- | --- | --- | --- |
| ASV000001 | *Klebsiella pneumoniae* | *K. pneumoniae* | NR_036794.1 |
| ASV000001 | *Klebsiella pneumoniae* | *K. pneumoniae* | NR_112009.1 |
| ASV000001 | *Klebsiella pneumoniae* | *K. pneumoniae* | NR_113240.1 |
| ASV000001 | *Klebsiella pneumoniae* | *K. pneumoniae* | NR_113702.1 |
| ASV000001 | *Klebsiella pneumoniae* | *K. pneumoniae* | NR_114506.1 |
| ASV000001 | *Klebsiella pneumoniae* | *K. pneumoniae* | NR_114715.1 |
| ASV000001 | *Klebsiella pneumoniae* | *K. pneumoniae* | NR_117682.1 |
| ASV000001 | *Klebsiella pneumoniae* | *K. pneumoniae* | NR_117683.1 |
| ASV000001 | *Klebsiella pneumoniae* | *K. pneumoniae* | NR_117684.1 |
| ASV000001 | *Klebsiella pneumoniae* | *K. pneumoniae* | NR_117685.1 |
| ASV000001 | *Klebsiella pneumoniae* | *K. pneumoniae* | NR_117686.1 |
| ASV000001 | *Klebsiella pneumoniae* | *K. pneumoniae* | NR_119278.1 |
| ASV000001 | *Klebsiella pneumoniae* subsp. *rhinoscleromatis* | *K. pneumoniae* | NR_037084.1 |
| ASV000001 | *Klebsiella pneumoniae* subsp. *rhinoscleromatis* ATCC 13884 | *K. pneumoniae* | NR_114507.1 |
| ASV000001 | *Klebsiella quasipneumoniae* subsp. *quasipneumoniae* | *K. pneumoniae* | NR_134062.1 |
| ASV000001 | *Klebsiella quasipneumoniae* subsp. *similipneumoniae* | *K. pneumoniae* | NR_134063.1 |
| ASV000001 | *Klebsiella variicola* | *K. pneumoniae* | NR_025635.1 |
| ASV000001 | *Klebsiella huaxiensis* | *K. oxytoca* | NR_171417.1 |
| ASV000001 | *Klebsiella aerogenes* | NA | NR_024643.1 |
| ASV000001 | *Klebsiella aerogenes* | NA | NR_113614.1 |
| ASV000001 | *Klebsiella aerogenes* | NA | NR_114737.1 |
| ASV000001 | *Klebsiella aerogenes* | NA | NR_118556.1 |
| ASV000001 | *Klebsiella aerogenes* KCTC 2190 | NA | NR_102493.2 |
| ASV000019 | *Klebsiella grimontii* | *K. oxytoca* | NR_159317.1 |
| ASV000019 | *Klebsiella michiganensis* | *K. oxytoca* | NR_118335.1 |
| ASV000019 | *Klebsiella oxytoca* | *K. oxytoca* | NR_041749.1 |
| ASV000019 | *Klebsiella oxytoca* | *K. oxytoca* | NR_112010.1 |
| ASV000019 | *Klebsiella oxytoca* | *K. oxytoca* | NR_113341.1 |
| ASV000019 | *Klebsiella oxytoca* | *K. oxytoca* | NR_114152.1 |
| ASV000019 | *Klebsiella oxytoca* | *K. oxytoca* | NR_118853.1 |
| ASV000019 | *Klebsiella oxytoca* | *K. oxytoca* | NR_119277.1 |

\*Only alignments with 100% identity are shown

**Table S3.** Clinical variables included in machine learning models*

| Variable |  | Case (N = 83) | Control (N = 147) |
| --- | --- | --- | --- |
| depression | yes | 29 (34.9%) | 39 (26.5%) |
|  | no | 54 (65.1%) | 108 (73.5%) |
|  | missing | 0 (0%) | 0 (0%) |
| prior diuretic | yes | 30 (36.1%) | 35 (23.8%) |
|  | no | 53 (63.9%) | 112 (76.2%) |
|  | missing | 0 (0%) | 0 (0%) |
| prior vitamin D | yes | 18 (21.7%) | 18 (12.2%) |
|  | no | 65 (78.3%) | 129 (87.8%) |
|  | missing | 0 (0%) | 0 (0%) |
| prior vasopressin blocker | yes | 19 (22.9%) | 14 (9.5%) |
|  | no | 64 (77.1%) | 133 (90.5%) |
|  | missing | 0 (0%) | 0 (0%) |
| albumin < 2.5 g/dL | yes | 34 (41%) | 34 (23.1%) |
|  | no | 46 (55.4%) | 106 (72.1%) |
|  | missing | 3 (3.6%) | 7 (4.8%) |
| high risk antibiotics | yes | 30 (36.1%) | 30 (20.4%) |
|  | no | 53 (63.9%) | 117 (79.6%) |
|  | missing | 0 (0%) | 0 (0%) |
| weighted elixhauser | mean ± SD | 22.4 ± 11.5 | 19.3 ± 12.5 |
| alcohol abuse | yes | 5 (6%) | 17 (11.6%) |
|  | no | 78 (94%) | 130 (88.4%) |
|  | missing | 0 (0%) | 0 (0%) |
| blood loss anemia | yes | 18 (21.7%) | 17 (11.6%) |
|  | no | 65 (78.3%) | 130 (88.4%) |
|  | missing | 0 (0%) | 0 (0%) |
| cardiac arrhythmias | yes | 50 (60.2%) | 83 (56.5%) |
|  | no | 33 (39.8%) | 64 (43.5%) |
|  | missing | 0 (0%) | 0 (0%) |
| chronic pulmonary disease | yes | 26 (31.3%) | 46 (31.3%) |
|  | no | 57 (68.7%) | 101 (68.7%) |
|  | missing | 0 (0%) | 0 (0%) |
| coagulopathy | yes | 36 (43.4%) | 53 (36.1%) |
|  | no | 47 (56.6%) | 94 (63.9%) |
|  | missing | 0 (0%) | 0 (0%) |
| congestive heart failure | yes | 28 (33.7%) | 47 (32%) |
|  | no | 55 (66.3%) | 100 (68%) |
|  | missing | 0 (0%) | 0 (0%) |
| deficiency anemia | yes | 11 (13.3%) | 20 (13.6%) |
|  | no | 72 (86.7%) | 127 (86.4%) |
|  | missing | 0 (0%) | 0 (0%) |
| complicated diabetes | yes | 15 (18.1%) | 21 (14.3%) |
|  | no | 68 (81.9%) | 126 (85.7%) |
|  | missing | 0 (0%) | 0 (0%) |
| uncomplicated diabetes | yes | 27 (32.5%) | 36 (24.5%) |
|  | no | 56 (67.5%) | 111 (75.5%) |
|  | missing | 0 (0%) | 0 (0%) |
| drug abuse | yes | 6 (7.2%) | 8 (5.4%) |
|  | no | 77 (92.8%) | 139 (94.6%) |
|  | missing | 0 (0%) | 0 (0%) |
| fluid electrolyte disorders | yes | 57 (68.7%) | 99 (67.3%) |
|  | no | 26 (31.3%) | 48 (32.7%) |
|  | missing | 0 (0%) | 0 (0%) |
| complicated hypertension | yes | 33 (39.8%) | 48 (32.7%) |
|  | no | 50 (60.2%) | 99 (67.3%) |
|  | missing | 0 (0%) | 0 (0%) |
| uncomplicated hypertension | yes | 35 (42.2%) | 86 (58.5%) |
|  | no | 48 (57.8%) | 61 (41.5%) |
|  | missing | 0 (0%) | 0 (0%) |
| hypothyroidism | yes | 15 (18.1%) | 16 (10.9%) |
|  | no | 68 (81.9%) | 131 (89.1%) |
|  | missing | 0 (0%) | 0 (0%) |
| liver disease | yes | 19 (22.9%) | 33 (22.4%) |
|  | no | 64 (77.1%) | 114 (77.6%) |
|  | missing | 0 (0%) | 0 (0%) |
| lymphoma | yes | 8 (9.6%) | 19 (12.9%) |
|  | no | 75 (90.4%) | 128 (87.1%) |
|  | missing | 0 (0%) | 0 (0%) |
| metastatic cancer | yes | 15 (18.1%) | 22 (15%) |
|  | no | 68 (81.9%) | 125 (85%) |
|  | missing | 0 (0%) | 0 (0%) |
| obesity | yes | 26 (31.3%) | 39 (26.5%) |
|  | no | 57 (68.7%) | 108 (73.5%) |
|  | missing | 0 (0%) | 0 (0%) |
| other neurological disorders | yes | 24 (28.9%) | 23 (15.6%) |
|  | no | 59 (71.1%) | 124 (84.4%) |
|  | missing | 0 (0%) | 0 (0%) |
| paralysis | yes | 6 (7.2%) | 4 (2.7%) |
|  | no | 77 (92.8%) | 143 (97.3%) |
|  | missing | 0 (0%) | 0 (0%) |
| peptic ulcer disease excluding bleeding | yes | 5 (6%) | 6 (4.1%) |
|  | no | 78 (94%) | 141 (95.9%) |
|  | missing | 0 (0%) | 0 (0%) |
| peripheral vascular disorders | yes | 17 (20.5%) | 41 (27.9%) |
|  | no | 66 (79.5%) | 106 (72.1%) |
|  | missing | 0 (0%) | 0 (0%) |
| psychoses | yes | 4 (4.8%) | 4 (2.7%) |
|  | no | 79 (95.2%) | 143 (97.3%) |
|  | missing | 0 (0%) | 0 (0%) |
| pulmonary circulation disorders | yes | 14 (16.9%) | 33 (22.4%) |
|  | no | 69 (83.1%) | 114 (77.6%) |
|  | missing | 0 (0%) | 0 (0%) |
| renal failure | yes | 26 (31.3%) | 35 (23.8%) |
|  | no | 57 (68.7%) | 112 (76.2%) |
|  | missing | 0 (0%) | 0 (0%) |
| rheumatoid arthritis collagen vascular diseases | yes | 8 (9.6%) | 10 (6.8%) |
|  | no | 75 (90.4%) | 137 (93.2%) |
|  | missing | 0 (0%) | 0 (0%) |
| solid tumor without metastasis | yes | 26 (31.3%) | 28 (19%) |
|  | no | 57 (68.7%) | 119 (81%) |
|  | missing | 0 (0%) | 0 (0%) |
| valvular disease | yes | 8 (9.6%) | 33 (22.4%) |
|  | no | 75 (90.4%) | 114 (77.6%) |
|  | missing | 0 (0%) | 0 (0%) |
| weight loss | yes | 45 (54.2%) | 49 (33.3%) |
|  | no | 38 (45.8%) | 98 (66.7%) |
|  | missing | 0 (0%) | 0 (0%) |
| urinary catheter | yes | 63 (75.9%) | 88 (59.9%) |
|  | no | 20 (24.1%) | 59 (40.1%) |
|  | missing | 0 (0%) | 0 (0%) |
| feeding tube | yes | 43 (51.8%) | 50 (34%) |
|  | no | 40 (48.2%) | 97 (66%) |
|  | missing | 0 (0%) | 0 (0%) |
| ventilator | yes | 38 (45.8%) | 67 (45.6%) |
|  | no | 45 (54.2%) | 80 (54.4%) |
|  | missing | 0 (0%) | 0 (0%) |
| central line | yes | 54 (65.1%) | 89 (60.5%) |
|  | no | 29 (34.9%) | 58 (39.5%) |
|  | missing | 0 (0%) | 0 (0%) |
| diabetes | yes | 30 (36.1%) | 41 (27.9%) |
|  | no | 53 (63.9%) | 106 (72.1%) |
|  | missing | 0 (0%) | 0 (0%) |
| hypertension | yes | 51 (61.4%) | 100 (68%) |
|  | no | 32 (38.6%) | 47 (32%) |
|  | missing | 0 (0%) | 0 (0%) |
| prior immunosuppressor | yes | 6 (7.2%) | 10 (6.8%) |
|  | no | 77 (92.8%) | 137 (93.2%) |
|  | missing | 0 (0%) | 0 (0%) |
| prior insulin | yes | 24 (28.9%) | 33 (22.4%) |
|  | no | 59 (71.1%) | 114 (77.6%) |
|  | missing | 0 (0%) | 0 (0%) |
| prior hypoglycemics | yes | 0 (0%) | 2 (1.4%) |
|  | no | 83 (100%) | 145 (98.6%) |
|  | missing | 0 (0%) | 0 (0%) |
| prior proton pump inhibitors | yes | 29 (34.9%) | 40 (27.2%) |
|  | no | 54 (65.1%) | 107 (72.8%) |
|  | missing | 0 (0%) | 0 (0%) |
| prior immunoglobulin | yes | 2 (2.4%) | 1 (0.7%) |
|  | no | 81 (97.6%) | 146 (99.3%) |
|  | missing | 0 (0%) | 0 (0%) |
| prior dialysis | yes | 1 (1.2%) | 0 (0%) |
|  | no | 82 (98.8%) | 147 (100%) |
|  | missing | 0 (0%) | 0 (0%) |
| prior nicotine | yes | 1 (1.2%) | 6 (4.1%) |
|  | no | 82 (98.8%) | 141 (95.9%) |
|  | missing | 0 (0%) | 0 (0%) |
| prior angiotensin blocker | yes | 0 (0%) | 8 (5.4%) |
|  | no | 83 (100%) | 139 (94.6%) |
|  | missing | 0 (0%) | 0 (0%) |
| prior antidepressant antipsychotic | yes | 22 (26.5%) | 30 (20.4%) |
|  | no | 61 (73.5%) | 117 (79.6%) |
|  | missing | 0 (0%) | 0 (0%) |
| prior histamine antagonists | yes | 16 (19.3%) | 29 (19.7%) |
|  | no | 67 (80.7%) | 118 (80.3%) |
|  | missing | 0 (0%) | 0 (0%) |
| prior antituberculars | yes | 1 (1.2%) | 1 (0.7%) |
|  | no | 82 (98.8%) | 146 (99.3%) |
|  | missing | 0 (0%) | 0 (0%) |
| prior clindamycin | yes | 3 (3.6%) | 1 (0.7%) |
|  | no | 80 (96.4%) | 146 (99.3%) |
|  | missing | 0 (0%) | 0 (0%) |
| prior cephalosporin | yes | 17 (20.5%) | 20 (13.6%) |
|  | no | 66 (79.5%) | 127 (86.4%) |
|  | missing | 0 (0%) | 0 (0%) |
| prior penicillin | yes | 24 (28.9%) | 26 (17.7%) |
|  | no | 59 (71.1%) | 121 (82.3%) |
|  | missing | 0 (0%) | 0 (0%) |
| prior quinolone | yes | 5 (6%) | 5 (3.4%) |
|  | no | 78 (94%) | 142 (96.6%) |
|  | missing | 0 (0%) | 0 (0%) |
| prior carbapenem | yes | 9 (10.8%) | 3 (2%) |
|  | no | 74 (89.2%) | 144 (98%) |
|  | missing | 0 (0%) | 0 (0%) |
| prior monobactam | yes | 3 (3.6%) | 1 (0.7%) |
|  | no | 80 (96.4%) | 146 (99.3%) |
|  | missing | 0 (0%) | 0 (0%) |
| prior aminoglycoside | yes | 14 (16.9%) | 6 (4.1%) |
|  | no | 69 (83.1%) | 141 (95.9%) |
|  | missing | 0 (0%) | 0 (0%) |
| prior macrolide | yes | 6 (7.2%) | 4 (2.7%) |
|  | no | 77 (92.8%) | 143 (97.3%) |
|  | missing | 0 (0%) | 0 (0%) |
| prior tetracycline | yes | 2 (2.4%) | 3 (2%) |
|  | no | 81 (97.6%) | 144 (98%) |
|  | missing | 0 (0%) | 0 (0%) |
| prior daptomycin | yes | 0 (0%) | 0 (0%) |
|  | no | 83 (100%) | 147 (100%) |
|  | missing | 0 (0%) | 0 (0%) |
| prior rifamycin | yes | 3 (3.6%) | 5 (3.4%) |
|  | no | 80 (96.4%) | 142 (96.6%) |
|  | missing | 0 (0%) | 0 (0%) |
| prior polymyxin | yes | 0 (0%) | 0 (0%) |
|  | no | 83 (100%) | 147 (100%) |
|  | missing | 0 (0%) | 0 (0%) |
| prior fosfomycin | yes | 1 (1.2%) | 0 (0%) |
|  | no | 82 (98.8%) | 147 (100%) |
|  | missing | 0 (0%) | 0 (0%) |
| prior nitrofurantoin | yes | 1 (1.2%) | 2 (1.4%) |
|  | no | 82 (98.8%) | 145 (98.6%) |
|  | missing | 0 (0%) | 0 (0%) |
| prior methotrexate | yes | 0 (0%) | 2 (1.4%) |
|  | no | 83 (100%) | 145 (98.6%) |
|  | missing | 0 (0%) | 0 (0%) |
| prior sulfonamide | yes | 4 (4.8%) | 1 (0.7%) |
|  | no | 79 (95.2%) | 146 (99.3%) |
|  | missing | 0 (0%) | 0 (0%) |
| prior 3rd or 4th generation cephalosporin | yes | 13 (15.7%) | 10 (6.8%) |
|  | no | 70 (84.3%) | 137 (93.2%) |
|  | missing | 0 (0%) | 0 (0%) |
| prior combo betalactam | yes | 24 (28.9%) | 23 (15.6%) |
|  | no | 59 (71.1%) | 124 (84.4%) |
|  | missing | 0 (0%) | 0 (0%) |
| prior linezolid | yes | 4 (4.8%) | 1 (0.7%) |
|  | no | 79 (95.2%) | 146 (99.3%) |
|  | missing | 0 (0%) | 0 (0%) |
| hemoglobin g/dL | mean ± SD | 7.4 ± 1.8 | 8.0 ± 2.1 |
| creatinine mg/dL | mean ± SD | 0.75 ± 0.48 | 0.78 ± 0.50 |
| albumin g/dL | mean ± SD | 2.5 ± 0.71 | 2.8 ± 0.72 |
| protein g/dL | mean ± SD | 4.8 ± 0.95 | 5.0 ± 0.93 |
| new ventilator | yes | 35 (42.2%) | 66 (44.9%) |
|  | no | 48 (57.8%) | 81 (55.1%) |
|  | missing | 0 (0%) | 0 (0%) |
| new urinary catheter | yes | 60 (72.3%) | 84 (57.1%) |
|  | no | 23 (27.7%) | 63 (42.9%) |
|  | missing | 0 (0%) | 0 (0%) |
| new feed tube | yes | 7 (8.4%) | 2 (1.4%) |
|  | no | 76 (91.6%) | 145 (98.6%) |
|  | missing | 0 (0%) | 0 (0%) |
| new central line | yes | 34 (41%) | 69 (46.9%) |
|  | no | 49 (59%) | 78 (53.1%) |
|  | missing | 0 (0%) | 0 (0%) |

\*Non-missing data in Table 1 were also included in machine learning models

